# COVI-Prim survey: Challenges for Austrian and German general practitioners during initial phase of COVID-19

**DOI:** 10.1101/2020.11.24.20237743

**Authors:** Andrea Siebenhofer, Sebastian Huter, Alexander Avian, Karola Mergenthal, Dagmar Schaffler-Schaden, Ulrike Spary-Kainz, Herbert Bachler, Maria Flamm

## Abstract

**Background:** Coronavirus disease 2019 (COVID-19) represents a significant challenge to health care systems around the world. A well-functioning primary care system is crucial in epidemic situations as it plays an important role in the development of a system-wide response.

**Methods:** 2,187 Austrian and German GPs answered an internet suvey on preparedness, testing, staff protection, perception of risk, self-confidence, a decrease in the number of patient contacts, and efforts to control the spread of the virus in the practice during the early phase of the COVID-pandemic (3rd to 30th April).

**Results:** The completion rate of the questionnaire was high (90.9%). GPs gave low ratings to their preparedness for a pandemic, testing of suspected cases and efforts to protect staff. The provision of information to GPs and the perception of risk were rated as moderate. On the other hand, the participants rated their self-confidence, a decrease in patient contacts and their efforts to control the spread of the disease highly.

**Conclusion:** Primary care is an important resource for dealing with a pandemic like COVID-19. The workforce is confident and willing to take an active role, but needs to be provided with the appropriate surrounding conditions. This will require that certain conditions are met.

**Registration:** Trial registration at the German Clinical Trials Register: DRKS00021231

**Primary Funding Source:** The study was financed by the cooperating University Institutes without any external financial support.

## Introduction

Coronavirus disease 2019 (COVID-19) represents a significant challenge to health care systems around the world. Although implications for the hospital and intensive care sector are generally focused on, a comprehensive approach to managing the COVID-19 pandemic should also involve primary care, as it is usually the point-of-first-contact, regardless of patients’ health concerns [1, 2]. In a pandemic, it is therefore particularly important that primary care is in a position to provide the continuous care that is needed, especially when other parts of the system are overwhelmed [3].

Primary care professionals represent the first point of contact in health care systems and are therefore in a vulnerable position. With sometimes insufficient information, they must deal with a dilemma between caring for potentially infectious patients [4], while protecting themselves and those around them from contracting the disease [5, 6]. Previous studies have emphasized the need to include general practitioners in preparedness planning and in supplying them with the personal protective equipment (PPE) they require to quickly adapt to highly dynamic epidemiological developments [7, 8]. While scenarios comparable to the COVID-19 pandemic have been simulated [9], national response plans in many countries still tend to neglect the primary care sector [10]. Furthermore, primary care in Austria and Germany is mostly delivered in small, decentralized units run by self-employed general practitioners (GPs), which may hinder a rapid and coordinated pandemic response [11].

Neither Germany nor Austria have yet exhausted their intensive care capacities and have managed to keep infection numbers under control [12, 13]. Nevertheless, it remains unclear how long the COVID-19 pandemic will last. Primary care will likely have to deal with recurring waves of infections, at least in certain regions, especially since dealing with viral infections is part of the daily business of general practice [14].

The aim of this study is to investigate the role played by GPs in the early phase of the COVID-19 pandemic, the specific challenges faced by them, their concerns and the strategies they have developed to cope with the pandemic. Potential deficiencies as well as regional differences (country-specific, setting, urbanity) are analyzed.

## METHODS

This manuscript was prepared in accordance with the CHERRIES criteria [15] (Supporting Information A-13). COVI-Prim-*Start* is part of the international COVI-Prim project [16]. Since this is the first publication to emerge from the project, the methods and design of the study are described in detail in the Supplement.

### Questionnaire development

To create a basic pool of items for the COVI-Prim questionnaire, we searched the literature for studies investigating the role of general practice during pandemics. Various topics, which had been partially grouped in topic areas in the literature, were identified. New topic areas were created for topics that did not belong in those found in the literature. Based on the literature review, semi-structured telephone interviews were carried out with GPs. The results were recorded using keywords and evaluated in terms of content and topic. New topics were identified in the first series of interviews (n = 9). A second series (n = 5) revealed no new topics, so we assumed that all relevant topics had been included. Based on these results, a questionnaire was developed that aimed to take all aspects into consideration, while being short enough to ensure a high response rate. The questionnaire was checked for comprehensibility by five GPs.

### Structure of the questionnaire

This analysis contains eight demographic items, 48 closed items (response scales: yes/no, yes/probably yes/probably no/no, very low/low/moderate/high/very high) and two items requiring GPs to provide exact numbers (e.g. “How many COVID-19 tests did you perform last week?”). The full questionnaire development is explained in the Supplement. The items not used in this paper will be analyzed in the longitudinal arm of the COVI-Prim study. Out of the 48 items used in this analysis eight factors were calculated. Reflecting the items contained within them, the factors were named as follows: (1) preparedness for a pandemic, (2) testing suspected cases, (3) protection of staff, (4) provision of information to GP, (5) perception of risk, (6) self-confidence, (7) decrease in number of patient contacts, (8) efforts to control the spread of the disease. Factor scores ranged from 0 – 10. The internal consistency (Cronbach’s Alpha) of these eight factors used in this analysis ranged from α = .48 to α = .85 (S1 Table).

### Survey

The questionnaire was transferred to LimeSurvey®. Invitations to GPs to respond to the questionnaire were sent out by participating universities in Austria (Graz, Salzburg, Innsbruck) and Germany (Frankfurt, Bochum, Hanover, Marburg, Gießen, Dresden, Freiburg, LMU Munich, Muenster, Aachen) using their respective mailing lists. Local GP associations, the Association of General Practitioners in Bavaria, Lower-Saxony and Baden-Wuerttemberg, Austria, and the Austrian Forum for Primary Care (OEFOP) also invited their members to participate. In accordance with data protection regulations, the study team did not have direct access to mailing lists. As the lists probably overlapped, it is not possible to know precisely how many GPs were contacted or to calculate a response rate. At the beginning of the survey, participants received information about its length, the investigators, and the purpose of the study. After ending the survey, all data on the online platform was stored in SPSS files. GPs received no incentive to participate.

### Statistics

Baseline characteristics are presented as mean ±SD or median (min-max), as appropriate. Categorical variables are provided as absolute numbers and in percent. In the main analysis, environmental variables (country of survey: Germany vs. Austria; size of town of practice: < 5,000 vs. 5,000 - <20,000 vs. 20,000 - <100,000 vs. ≥100,000; type of practice: single-handed vs. not single handed) that may have influenced the responses were analyzed using General Linear Models. The main effects and all two-way interactions were therefore analyzed. Bonferroni correction was used to take account of multiple testing. Estimated means and 95% confidence intervals were used to present the results. For a better understanding of the results, responses to the items were also presented. In this presentation, the response categories “yes” and “probably yes” and the response categories “probably no” and “no” were combined. No statistical correction was carried out to adjust for non-representative samples.

### Ethics

The study protocol has been approved by the local ethics committee of Goethe University Frankfurt, Germany (20-619).

## RESULTS

### Demographics

The survey was answered by 2,187 Austrian and German GPs during the early phase of the COVID-19-pandemic (3^rd^ April to 30^th^ April). The majority of GPs were male (55.6%), practiced in a city with fewer than 20,000 inhabitants (59.4%) and had a single-handed practice (57.7%). Mean age of the GPs was 52.5 years (SD: 9.6). In the week prior to answering the questionnaire, 56.1% of the GPs (n = 1226) ordered at least one COVID-19 test. In total 13,520 tests were ordered. Of the 1,226 GPs that ordered COVID-19 tests, 41.0% (n = 503; 41 GPs did not answer the question on the test results) received positive results for 1,593 patients (12.1% of 13,139 tests; 12.1%). All demographic characteristics are provided in Table 1.

**Table 1.** Baseline demographics.

|  | All | Germany | Austria |
| --- | --- | --- | --- |
|  | n=2187 | n=1287 | n=900 |
| Age (years) | 52.2 ± 9.6 | 51.7 ± 9.5 | 53.8 ± 9.6 |
| Sex |  |  |  |
| male | 1217 (55.6%) | 673 (52.3%) | 544 (60.4%) |
| female | 965 (44.1%) | 609 (47.3%) | 356 (39.6%) |
| other | 5 (0.2%) | 5 (0.4%) | 0 (0.0%) |
| Size of town of practice |  |  |  |
| < 5,000 | 658 (30.1%) | 264 (20.5%) | 394 (43.8%) |
| 5,000 - <20,000 | 642 (29.4%) | 421 (32.7%) | 221 (24.6%) |
| 20,000 - <100,000 | 635 (16.1%) | 287 (22.3%) | 66 (7.3%) |
| ≥100,000 | 534 (24.4%) | 315 (24.5%) | 219 (24.3%) |
| Type of practice |  |  |  |
| single-handed | 1262 (57.7%) | 505 (39.2%) | 757 (84.1%) |
| not single-handed | 952 (42.3%) | 782 (60.8%) | 143 (15.9%) |
| Position in the practice |  |  |  |
| employed | 213 (9.7%) | 202 (15.7%) | 11 (1.2%) |
| owner | 1945 (88.9%) | 1080 (83.9%) | 865 (96.1%) |
| locum | 29 (1.3%) | 5 (0.4%) | 24 (2.7%) |
| Year practice was established | median: 2003<br>Range: 1975 - 2020 | 2005<br>1975 – 2020 | 2003<br>1975 - 2020 |
| GPs that ordered COVID-19 tests in previous 7 days |  |  |  |
| no | 760 (34.8%) | 289 (22.5%) | 471 (52.3%) |
| yes | 1226 (56.1%) | 916 (71.2%) | 310 (34.4%) |
| missing | 201 (9.2%) | 82 (6.4%) | 119 (13.2%) |
| GPs with patients with positive COVID-19 test results in previous 7 days (n = 1226) |  |  |  |
| no | 682 (55.6%) | 520 (56.8%) | 162 (52.3%) |
| yes | 503 (41.0%) | 368 (40.2%) | 135 (43.5%) |
| missing | 41 (3.3%) | 28 (3.1%) | 13 (4.1%) |

Of the 2,187 GPs, 1,989 (90.9%) rated enough items to be included in the analysis. The median time required to answer the questionnaire was 14.1 minutes (IQR: 10.5 – 20.2 minutes) in Austria and 13.4 minutes (IQR: 9.8 – 19.0) in Germany. The completion rate of the survey was 79.7% in Austria and 85.2% in Germany.

### Overall results

GPs gave low ratings to their preparedness for a pandemic (mean: 2.7; 95% CI: 2.5-2.8, n = 1989), testing of suspected cases (3.3, 95%CI 3.2-3.4) and efforts to protect staff (2.0 95%CI 1.9-2.1). The provision of information to GPs (4.3, 95%CI: 4.2-4.4) and the perception of risk (5.1 95%CI 4.9-5.2) were rated as moderate. On the other hand, the participants rated their self-confidence (7.7, 95%CI 7.5-7.8), a decrease in patient contacts (6.8, 95%CI 6.7-7.0) and their efforts to control the spread of the disease (7.3, 95%CI 7.2-7.4) highly.

### Pandemic preparedness

Looking back to the beginning of the pandemic, 88.2% of GPs said they did not have enough protective equipment and 91.4% stated that they did not receive sufficient information on how much protective equipment they needed. Furthermore, a substantial number of GPs did not know where to procure protective equipment (78.3%) and said their practice was not well prepared for the COVID-19 pandemic (77.2%).

### Testing of suspected cases

Of the participants, 92.5% agreed that GPs should decide which patients should undergo testing for COVID-19. The idea of a telephone hotline for the exclusive use of medical staff ordering COVID-19 tests was approved by 86.9% of respondents. Of the GPs, 83.6% rejected the idea that all suspected cases of COVID-19 should be sent directly to hospital to enable them to focus on other patients. Furthermore, a large number of GPs said too little testing is performed (71.9%) and that they did not have adequate access to tests at the beginning of the pandemic (71.0%).

### Decrease in patient contacts

Of the GPs, 95.2% had less contact to patients as a result of the pandemic. Of these, 71.9% said they had less workload at the time because many patients are avoiding coming to the practice.

### Information

Of the GPs, 71.4% said they had received insufficient information from public bodies. Before officially informing GPs of new developments, public authorities distributed important information to the general public via the media (70.9%).

### Self-confidence

Almost all the GPs said they knew what to do in suspected cases of COVID-19 (99.1%), and 82.1% were convinced they knew enough to provide optimal care for their patients during the pandemic.

### Efforts to control the spread of the virus in the practice

Almost all GPs tried to gain enough information from patients by phone beforehand to know whether they were dealing with a suspected case of COVID-19 (98.5%), and they took precautions to ensure that suspected cases did not come into contact with other patients in their practice (97.4%). Over 80% of GPs avoided treating patients with mild symptoms that were not clearly linked to suspected cases of COVID-19 in their practice and preferred to attend to them by phone or online (87.9%). The distribution of responses is given in S3 Table.

### Economic aspects

60.0% of GP were concerned about how the pandemic would affect their own and their employees’ economic prospects.

### Regional differences

Differences in the GP’s responses were found to depend on the country in which the survey was conducted and the size of the city in which the practice was located. Whether the practice was single-handed or not did not influence GP’s responses. Furthermore, no interactions between observed variables were significant.

Compared to Austrian GPs, German GPs rated their self-confidence lower (Germany: 7.5 95%CI: 7.4-7.6 vs. Austria: 7.8 95%CI: 7.6-8.0; p = .009), as they did their efforts to control the spread of SARS-CoV-2 (Germany: 7.1 95%CI: 7.0-7.2 vs. Austria: 7.5 95%CI: 7.3-7.6; p = .001). However, they rated their testing of suspected cases higher (Germany: 4.0 95%CI: 3.9-4.2 vs. Austria: 2.5 95%CI: 2.3-2.7; p = .009) and were more likely to say the number of patient contacts had decreased (Germany: 7.1 95%CI: 7.0-7.1 vs. Austria: 6.6 95%CI: 6.4-6.8; p < .001) (Table 2, Fig 1). Looking at single items, the biggest difference between German and Austrian GPs was found in testing, with 62.8% of German GPs saying too little testing was carried out, compared to 84.9% of Austrian GPs, and 42.4% of German GPs saying they had adequate access to tests at the beginning of the pandemic, compared to 9.7% of Austrian GPs.

**Table 2.** Mean and 95%CI for each factor of the evaluation of the pandemic for the whole group and subgroups. Significant differences are in bold. (Scale values range from 0 – 10)

|  |  | Type of practice<br>(single-handed) |  | Country of survey |  | City size |  |  |  |
| --- | --- | --- | --- | --- | --- | --- | --- | --- | --- |
|  | overall |  |  |  |  |  |  |  |  |
|  |  | yes | no | Austria | Germany | <5,000 | 5,000 –<br><20,000 | 20,000<br>–<br><100,000 | ≥ 100,000 |
| Preparedness for a pandemic | 2.7 (2.5-2.8) | 2.5 (2.4-2.7) | 2.8 (2.6-3.0) | 2.6 (2.4-2.8) | 2.7 (2.6-2.9) | 2.7 (2.5-2.8) | 2.5 (2.4-2.7) | 2.8 (2.5-3.1) | 2.6 (2.4-2.8) |
| Testing of suspected cases | 3.3 (3.2-3.4) | 3.2 (3.1-3.3) | 3.3 (3.2-3.5) | 2.5 (2.3-2.7) | 4.0* (3.9-4.2) | 3.2 (3.1-3.4) | 3.3 (3.1-3.4) | 3.4 (3.1-3.6) | 3.2 (3.0-3.4) |
| Protection of staff | 2.0 (1.9-2.1) | 1.8 (1.7-2.0) | 2.2 (2.0-2.4) | 2.1 (1.9-2.4) | 1.9 (1.7-2.0) | 2.0 (1.8-2.2) | 2.1 (1.9-2.3) | 1.9 (1.6-2.2) | 2.0 (1.8-2.2) |
| Provision of information to GPs | 4.3 (4.2-4.4) | 4.3 (4.2-4.5) | 4.3 (4.1-4.5) | 4.2 (3.9-4.4) | 4.5 (4.3-4.6) | 4.4 (4.2-4.6) | 4.3 (4.1-4.5) | 4.4 (4.1-4.7) | 4.2 (3.9-4.4) |
| Perception of risk | 5.1 (4.9-5.2) | 5.0 (4.8-5.2) | 5.1 (4.9-5.4) | 4.8 (4.6-5.1) | 5.3 (5.1-5.4) | 5.1 (4.9-5.4) | 5.0 (4.8-5.3) | 5.1 (4.7-5.5) | 5.0 (4.8-5.3) |
| Self-confidence | 7.7 (7.5-7.8) | 7.7 (7.5-7.7) | 7.6 (7.5-7.8) | 7.8 (7.6-8.0) | 7.5* (7.4-7.6) | 7.6<sup>†</sup> (7.5-7.8) | 7.6 (7.5-7.8) | 8.0<sup>†</sup> (7.7-8.2) | 7.4<sup>‡,§</sup> (7.2-7.5) |
| Decrease in number of patient contacts | 6.8 (6.7-7.0) | 6.9 (6.8-7.1) | 6.8 (6.5-7.0) | 6.6 (6.4-6.8) | 7.1* (7.0-7.2) | 6.7 (6.5-6.9) | 6.7 (6.6-6.9) | 7.0 (6.7-7.3) | 6.9 (6.7-7.1) |
| Efforts to control the spread of<br>the disease in the practice | 7.3 (7.2-7.4) | 7.2 (7.1-7.3) | 7.4 (7.3-7.6) | 7.5 (7.3-7.6) | 7.1* (7.0-7.2) | 7.3 (7.2-7.5) | 7.3 (7.2-7.5) | 7.2 (7.0-7.4) | 7.3 (7.2-7.5) |

\* Comparison Austria vs. Germany, p <.05
<sup>†</sup> ... Variable city size: Post Hoc comparison to ≥ 100,000, p <.05 (Bonferroni corrected)
<sup>‡</sup> ... Variable city size: Post Hoc comparison to <5,000, p <.05 (Bonferroni corrected)
<sup>§</sup> ... Variable city size: Post Hoc comparison to 20,000 - <100,000, p <.05 (Bonferroni corrected)

**Fig 1.**
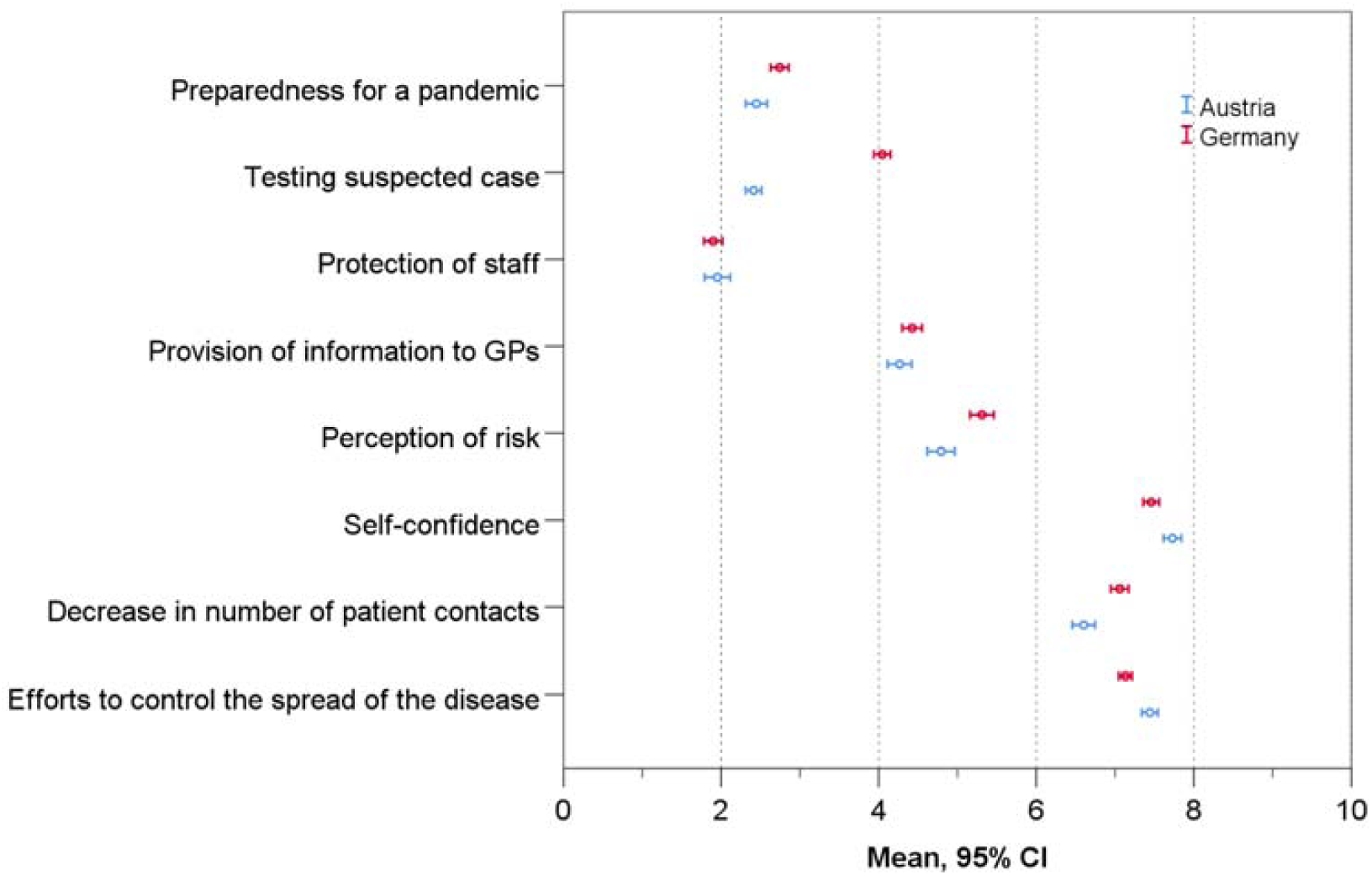
Differences between German and Austrian GPs in their evaluation of the pandemic (Austria: n = 900; Germany: n = 1287)

GPs in cities with 100,000 inhabitants or more rated their self-confidence lower than GPs in towns with fewer than 5,000 (p = .041) and towns with 20,000 – 100,000 (p <.001) inhabitants (Fig 2, Table 2). Analyzing the items used to calculate the self-confidence score, the largest difference can be observed in GPs’ conviction that their knowledge was sufficient to provide optimal care for their patients during the pandemic. While 87.1% of GPs in cities with 20,000-100,000 inhabitants were convinced, the number fell to 82.9% in cities with fewer than 5,000 inhabitants and to 79.0% in cities with 100,000 or more inhabitants.

**Fig 2.**
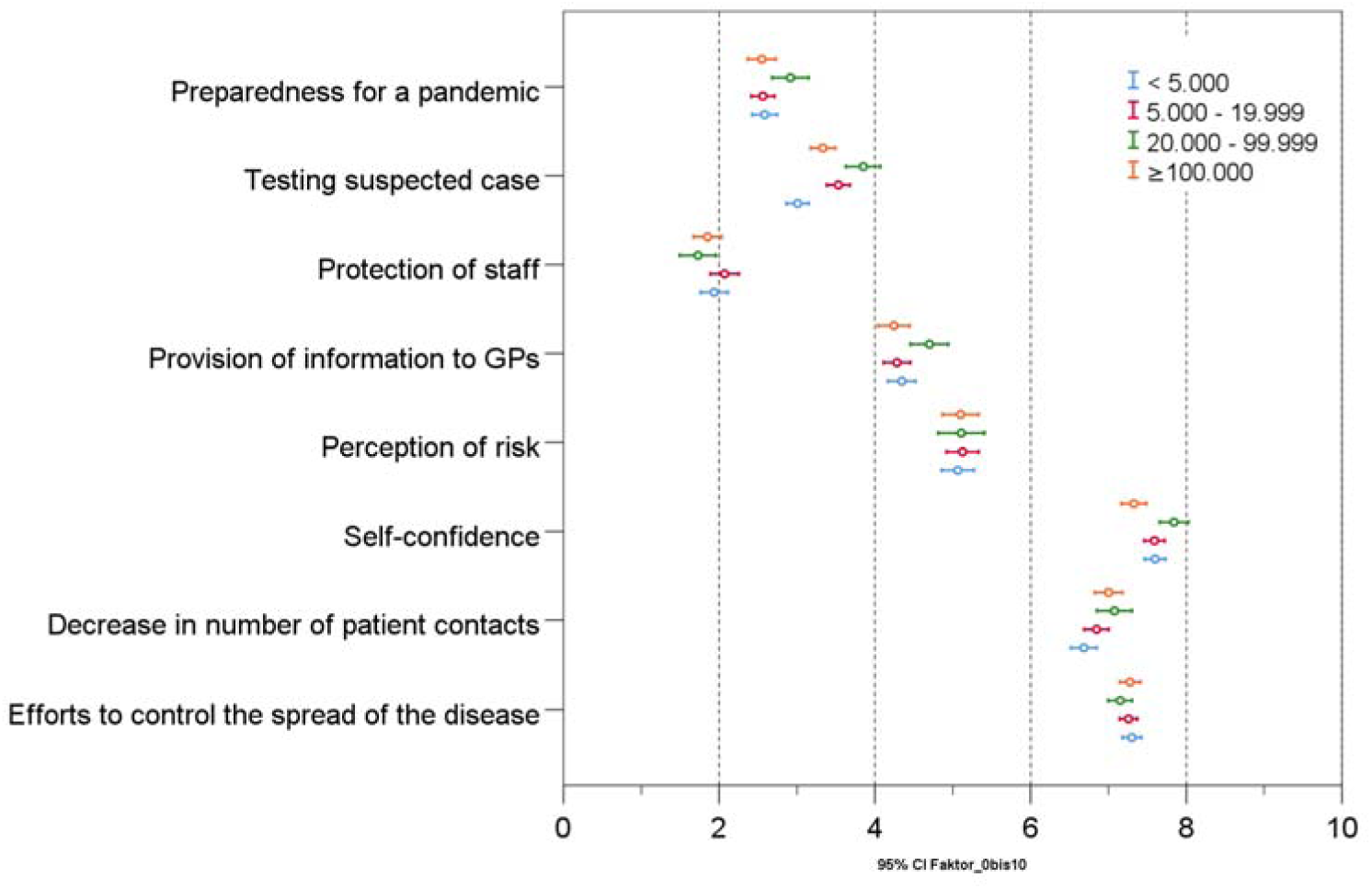
Differences in the evaluation of the pandemic of GPs with practices in cities of different sizes.

## DISCUSSION

Our survey covered the specific problems and experiences of more than two thousand general practitioners in Austria and Germany at the beginning of the COVID-19 pandemic. The high level of participation demonstrates the interest and concern of this group. In the early stages, GP practices were not well prepared and did not have enough protective equipment. GPs did not receive sufficient information from public stakeholders but were very active on informal digital networks involving their professional peer group. Overall, they had fewer patient contacts. A majority wanted to decide themselves whom to test, and to have a higher number of tests made available to GPs themselves. They were concerned about the economic outlook but they were generally self-confident in terms of dealing with suspected and confirmed cases of COVID-19. Considering its scale and abruptness, the reported lack of preparation for an event such as the COVID-19 pandemic is not surprising. Even though GPs immediately went to great lengths to procure enough protective equipment and to re-organize and adapt standard procedures in their practices, some – as in other countries – also had to work without sufficient PPE [17-19]. Since the availability of PPE is essential to ensuring the continuous and safe provision of care during a pandemic, it is critical to incorporate primary care practices in the procurement of PPE. Existing structures should support the development of a joint national response plan to ensure that primary care is adequately involved [10].

Although many of the challenges such as that mentioned above were observed internationally, some regional differences stand out. In particular, GPs in Austria were not initially involved in testing procedures. Instead, the population in Austria was encouraged to contact an official health hotline in case of symptoms or suspicion of infection. Hence, GPs were overlooked in their role as gatekeepers in primary care. For GPs, this is likely to have been particularly frustrating, as the vast majority are convinced they know how to manage patients with a suspected infection and are willing to do so.

Furthermore, in the current situation it is especially important to motivate primary care practitioners, as they are in the frontline in terms of contact with the community [3]. The role of the GP is to decide which patients need hospital care and to monitor others at home [20]. This is the only way to ensure that important resources in hospitals are not overburdened. Experts’ concerns that a significant number of patients may die or suffer harm due to delayed access to usual medical care [21, 22] are also important and are reflected in our survey. As noted above, the number of patients visiting primary care practices decreased during the COVID-19 pandemic. People had strict stay-at-home orders or were afraid of infection. However, a few weeks after the lockdown, there was widespread criticism that this may have led to significant collateral damage. Several recently published articles pointed out that fewer patients were diagnosed with serious medical conditions such as stroke [23], acute coronary syndrome [24], atrial fibrillation [25] and cancer [26]. Furthermore, the WHO warned that measures designed to slow the spread of the coronavirus might also delay vaccination programs and thereby speed up the spread of other vaccine-preventable diseases [27].

General practitioners are responsible for the population as a whole, and the COVID-19 pandemic affected everyone. While children usually only experience mild or asymptomatic disease symptoms [28], they are also strongly affected by social isolation. A lack of structure and support from schools can increase anxiety and potentially impact mental health [29]. Other vulnerable groups to consider are elderly people that are living alone and for whom the use of online communication systems is often not feasible, as well as those with mental health problems, or people living in poor socio-economic conditions. They are all part of the patient collective in a primary care setting. We therefore need strategies to avoid future collateral damage that ensure access to primary care, even at times of high infection rates. Possible solutions, such as the greater use of telemedicine appointments and triage for certain patient groups according to the severity and urgency of a consultation, are surveyed in our longitudinal study (see supporting information), for which the analysis is ongoing.

But telemedicine alone is not enough. About 60% of GPs reported financial and economic concerns. This suggests that existing remuneration mechanisms for primary care need to be adapted or amended during a pandemic. Basu et al. estimated that the losses to primary care practices resulting from the pandemic amounted to about 15 billion USD in the U.S. alone [30].While SARS-CoV-2 is certainly the most serious pandemic since the influenza pandemic of 1917-18 [31], it has not been the only one in recent years. The H1N1 virus in 2009 was also declared responsible for an influenza pandemic and resulted in widespread preparations. However, it had far less impact on the population than expected, and a specific vaccine and treatment was available early [32]. SARS-CoV-1 in 2003 resulted in a similar public health response in strongly affected regions like Toronto [33]. Many of the issues that arose during that outbreak are mirrored in this pandemic on a global scale and can be found in the results of our study. Such pandemics, as well as seasonal influenza epidemics, lead to a surge in hospital bed demand and primary care consultations [34]. The COVID-19 pandemic is somewhat different because a strong focus was placed on saving health care resources in countries that had time to prepare before the need for them had arisen.

Our study has some limitations. Firstly, the questionnaire was developed in a very short time so that it could be delivered when the situation was most acute. Even though we tried to include all relevant topics, some issues may have been missed. Secondly, we could not calculate the response rate because a systematic area-wide survey was not possible in the time frame we permitted ourselves. However, the number of responses far exceeded our expectations, especially considering the difficulties that are usually encountered in recruiting GPs for research projects [35]. In addition, the questionnaire was completed by a very high percentage of participants. Thirdly, the recruitment process through regional networks and professional associations led to the heterogeneous selection of participants, which may have limited representativeness. One further limitation is that our survey was only carried out among GPs and did not involve other team members from the primary care setting.

Primary care is an important and vital resource for dealing with a pandemic like COVID-19. The workforce is confident and willing to take an active role, but needs to be given the opportunity and provided with the necessary conditions to do so. As GPs work on the frontline, they should be adequately supported, both in terms of the provision of protective equipment and financial security during the active phase of the pandemic. To ensure a quick and effective response to any new crisis, general practitioners in primary care should be involved in a national coordinated strategy that includes all relevant parties.

## Data Availability

The authors have uploaded an additional minimal data set to Data Archiving and Networked Services (DANS)

https://doi.org/10.17026/dans-z8u-unbv

## Acknowledgments

We would like to thank all participating general practitioners, and the institutions that were willing to send the link to our questionnaire to their network partners.

## Supporting information

COVI-Prim is an international project that plans to carry out regular surveys of GPs working in primary care during the COVID-19 pandemic in order to research their role in it, the specific challenges they face, and the strategies they have developed to deal with it (https://allgemeinmedizin.medunigraz.at/news/, http://www.allgemeinmedizin.uni-frankfurt.de/forschung1/covi_prim.html, https://www.pmu.ac.at/allgemeinmedizin.html). Potential deficiencies in care and possible obstacles such as a lack of stakeholder support are analyzed. An overview of the COVI-Prim project is provided in **Appendix Table 1**.

**Appendix Table 1.** COVI-Prim overview.

| Participating Countries | Start of project | Finalization / Translation of Questionnaire | Ethics approval | Start of the survey | End of the survey | Baseline survey | Longitudinal survey | Sub - Project |  |  |  |
| --- | --- | --- | --- | --- | --- | --- | --- | --- | --- | --- | --- |
|  |  |  |  |  |  |  |  | COVI-Prim-<br>*Start* | COVI-Prim-<br>*Flat* | COVI-Prim-<br>*Long* | COVI-Prim-<br>*Hot topics* |
| Australia | 8<sup>th</sup> April | 20<sup>th</sup> April | Yes | 8<sup>th</sup> May | 8<sup>th</sup> August | x | x |  | X | X |  |
| Austria | 20<sup>th</sup> March | 30<sup>th</sup> March | N.A. | 3<sup>rd</sup> April | 29<sup>th</sup> May | x | x | X | X | X | x |
| Germany | 20<sup>th</sup> March | 30<sup>th</sup> March | Yes | 3<sup>rd</sup> April | 27<sup>th</sup> May | x | x | X | X | X | x |
| Hungary | 7<sup>th</sup> May | 3<sup>rd</sup> June | N.A. | 5<sup>th</sup> June | 2<sup>nd</sup> July | x |  |  | X |  |  |
| Italy/German | 23<sup>rd</sup> April | German | N.A. | 23<sup>rd</sup> April | 6<sup>th</sup> May | x |  |  | x |  | x |
| Slovenia | 29<sup>th</sup> April | 15<sup>th</sup> May | Yes | 1<sup>st</sup> June | 6<sup>th</sup> July | x |  |  | x |  |  |
| Switzerland | 15<sup>th</sup> May | German/Italian<br>(31<sup>st</sup> May) | N.A. | 7<sup>th</sup> July | 4<sup>th</sup> August | x |  |  | x |  |  |

N.A. ...Not applicable

### Questionnaire development

To create a basic item pool for the COVI-Prim questionnaire, we searched the literature for studies investigating the role of general practice during past pandemics. The search revealed a number of topics, some of which had been grouped to form topic areas in the literature. New topic areas were created for topics that did not fit into one of these. Based on the literature review, semi-structured telephone interviews were carried out with GPs. The results were recorded in keywords and evaluated in terms of content and topic. After identifying new topics in a first series of surveys (n = 9), no further new topics were found in a second (n = 5). It was therefore assumed that all relevant topics had been identified.

**Appendix Fig 1.**
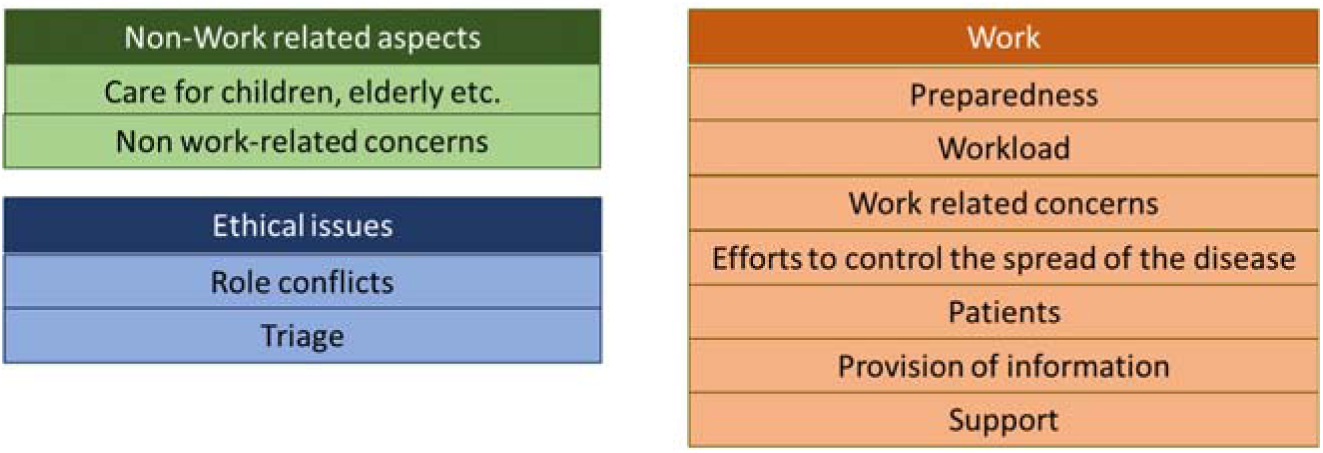
Topic areas according to literature review and interviews.

The literature and interviews revealed the following topic areas:

Based on this structure, a questionnaire was developed that aimed to take all aspects into consideration, while being short enough to ensure a high response rate. The questionnaire was checked for comprehensibility by five GPs.

A short version of the questionnaire was prepared for a longitudinal survey. The items with potentially time-sensitive content were selected for the short version, as we assumed responses to these items might change during the pandemic. At the end of the full version of the questionnaire, each respondent had the opportunity to give his/her active consent to participate in the longitudinal survey (every 1 – 2 weeks) by providing their e-mail address.

### Structure of the Questionnaires

The full questionnaire consisted of eight demographic items, 48 closed items (response scales: yes/no, yes/probably yes/probably no/no, very low/low/moderate/high/very high), three items requiring GPs to provide exact numbers (e.g. “How many COVID-19 tests did you perform last week?”), seven items requiring GPs to provide proportions (e.g. “How much of your overall working time was directly or indirectly linked to COVID-19?”), and five open-ended questions. The items in the questionnaire were grouped into seven sections: (1) demographic items, (2) preparedness at the beginning of the pandemic, (3) provision of information to GPs during the pandemic, (4) management of the pandemic by GPs, (5) personal worries, (6) personal emotions and (7) work content and burden of work. Overall, the questionnaire consisted of six pages.

To identify uncorrelated factors, exploratory factor analysis (VARIMAX rotation) was calculated for the following items: preparedness at the beginning of the pandemic, provision of information to GPs during the pandemic, management of the pandemic by GPs, and personal worries. To determine how many factors to retain, we applied Horn’s parallel analysis and the criterion of eigenvalue > 1. Difficult items, defined as items for which more than 90% of responses fell into one of the two extreme categories, were excluded from the analysis. Ten factors had eigenvalues >1. Based on Horn’s parallel analysis, the original high number of factors fell to eight, with each explaining 3.0% to 15.2% of the variance (total variance explained = 46.4%). After eliminating all items with double loadings (items loading on two factors within a range of .1) and the highest factor loadings ≤ .3, a version of the questionnaire with 39 items remained. No item had to be excluded because of too many responses in an extreme response category. One item was excluded because of a mismatch between the item and factor content. Each of the remaining 38 items was assigned to one of the eight factors, with the factors ultimately including three to seven items. Internal consistency (Cronbach’s Alpha) of these eight factors ranged from α = .48 to α = .85.

**Appendix Table 2.** Internal consistency of the factors used to evaluate the pandemic

|  | Cronbach's alpha |
| --- | --- |
| Perception of risk | .851 |
| Provision of information to GPs | .810 |
| Preparedness for a pandemic | .726 |
| Self-confidence | .593 |
| Testing suspected cases | .557 |
| Decrease in number of patient contacts | .567 |
| Efforts to control the spread of the disease | .483 |
| Protection of staff | .484 |

Reflecting the items contained within them, the factors were named as follows: (1) Preparedness for a pandemic, (2) Testing suspected cases, (3) Protection of staff, (4) Provision of information to GPs, (5) Perception of risk, (6) Self-confidence, (7) Decrease in number of patient contacts, (8) Efforts to control the spread of the virus in the practice. To calculate factor scores (*f*_*x*_), the mean score of the items was calculated for each scale. The resulting score, which ranged from 1 to 4, was linearly transformed to 0-10 for a better interpretability (*f*_*xneu*_ *= (f*_*x*_ *-1) * 3*^*-1*^ *\* 10*). Calculation of factor scores was only performed when fewer than 50% of items were missing. To evaluate the effect of calculating factor scores with missing values, factor scores calculated form a complete response set analyzed. For this purpose, within the complete response set, single responses were randomly deleted (response set with missing values) from the original set of responses. The factor scores derived from the original response set (complete response set) was correlated with the factor score derived from the response set with missing values. The correlation between factor scores calculated with missing values and the factor score without missing values was r = .943 (95%CI: .938 -.947) for a seven-item scale with one missing item, r = .880 (95%CI: .869 - .889) with two missing items and r = .799 (95%CI .793 - .813) with three missing items.

The <u>longitudinal questionnaire</u> consisted of 15 closed items, three items required GPs to provide exact numbers, seven items required them to provide proportions, and five were open-ended items (3 pages). Of these 15 closed items, the factors “perception of risk” and “decrease in number of patient contacts” could be calculated. To ensure the response rate was high every week, the German version of the longitudinal survey was extended to include “hot topics”. The choice of hot topic was selected on the basis of weekly responses to the open-ended question “What was the biggest challenge you had to face as a GP this week?” A topic was then chosen, according to the previous week’s answers. The chosen topics were: Provision of information (survey period: 10.4. – 16.4 2020; response rate: 39%), Telemedicine (survey period: 17.4. – 23.4 2020; response rate: 55%), “Overlooked” patient groups (survey period: 24.4. – 30.4 2020; response rate: 45%), Residents of nursing homes for the elderly (survey period: 1.5. – 7.5 2020; response rate: 44%), Economic consequences for GPs (survey period: 8.5. – 21.5 2020; response rate: 39%), Personal evaluation of the lockdown (survey period: 22.5. – 4.6 2020; response rate: 38%), Lessons learned (survey period: 5.6. – 18.6 2020; response rate: 39%), and Evaluation of the role of GPs during the pandemic (survey period: 19.6. – 2.7 2020; response rate: 30%). Based on the comments of the GPs, an expert group created and discussed the items (GPs, Psychologist).

In Australia, follow-up surveys were performed on 23^rd^ May (response rate: 20 out of 21 contacted GPs), 6^th^ June (response rate: 13 out of 15), 20^th^ June (response rate: 7 out of 10), 4^th^ July (response rate: 5 out of 5), 18^th^ July (response rate: 1 out of 3) and 1^st^ August (response rate: 0 out of 1%).

### Translation

The final baseline and longitudinal questionnaires were translated into English, Hungarian, Slovenian and Italian. The translation process for each language consisted of a translation (German to target language), followed by a back translation (target language to German). The back-translated version was compared to the original German version by a psychometrician, and the target language version was adapted where necessary.

### Survey

The long version of the questionnaire and the short version of the questionnaire were transferred to LimeSurvey in all five languages. Since the developmental process of the translated questionnaires took time, the German version was the only one to be distributed at the beginning of April 2020 (COVI-Prim-*Start*; responses: Germany: 3^rd^ April – 27^th^ May, Austria: 3^rd^ April – 29^th^ May). The open survey began in the second half of April in Italy (23^rd^ April – 6^th^ May), at the beginning of May in Australia (8^th^ May – 8^th^ August), at the beginning of June in Slovenia (1^st^ June – 6^th^ July) and Hungary (5^th^ June – 2^nd^ July), and at the beginning of July in Switzerland (7^th^ July – 4^th^ August) (Appendix Figure 2). Participation was voluntary and participants received no incentives.

**Appendix Fig 2.**
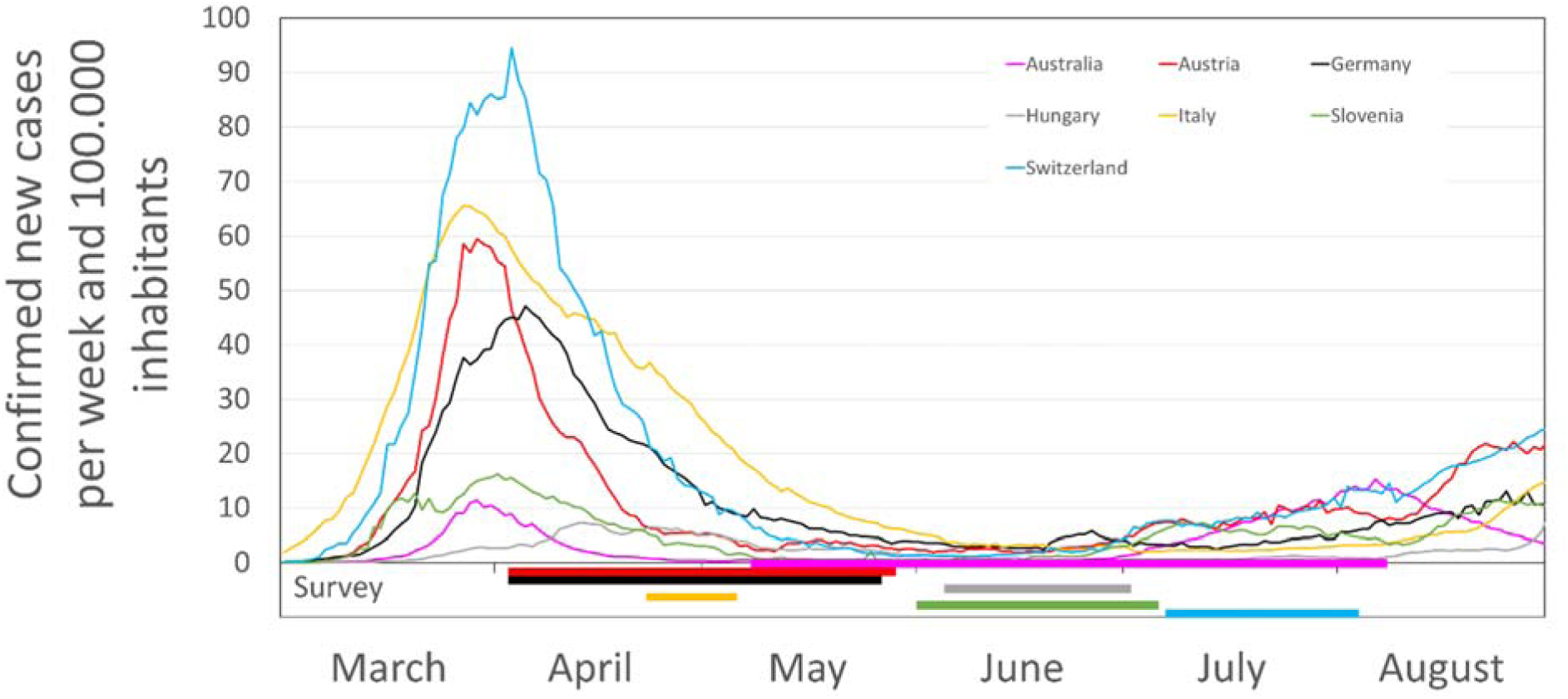
Confirmed new cases per week and 100,000 inhabitants from 1^st^ March until 31^st^ of July. timeline of survey in each country. (Source: https://covid19.who.int/table ; 1^st^ September 2020)

Invitations to respond to the questionnaire were sent out by participating universities (Australia: Bond University, Queensland; Austria: Graz, Salzburg, Innsbruck; Germany: Frankfurt, Bochum, Hannover, Marburg, Gießen, Dresden, Freiburg, LMU Munich, Muenster, Aachen; Slovenia: Maribor; Switzerland: Bern) to GPs in their mailing lists. Local general practice associations (Styrian, Tyrolean, Carinthian, Vienna Society for General Practice), the Association of General Practitioners in Bavaria, Lower-Saxony and Baden-Wuerttenmberg and Austria. Michael Kochen of DEGAM-Benefits and the Austrian Forum for Primary Care (OEFOP) also invited their members to participate. In accordance with data protection regulations, the study team did not have direct access to the mailing lists. As these lists are likely to overlap, it is not possible to know the exact number of contacted GPs. A calculation of response rate is therefore not possible. Items were presented in a non-randomized manner. Some items for the “hot topics” were presented adaptively (e.g. different items were presented depending on whether a GP said she or he was the owner of the practice). No completeness check was performed before submission of the questionnaires. Participants could not review and change their responses after they had completed a page and started to respond on the next. Neither cookies, IP checks nor log file analyses were used to identify multiple entries. Atypical timestamps were not used to delete questionnaires responses. At the beginning of the survey, participants were informed about the length of the survey, who the investigator was, and the purpose of the study. Furthermore they were informed about the management of their data (which data, where and how long they are stored, access to the data). Before participants could start to answer the items, they had to state, that they have read this information and gave consent. After ending the survey, all data on the online platform were stored in SPSS files. GPs were offered no incentive or reward for their participation. The median time required to answer the questionnaire was 11:00 minutes (interquartile range: 7:36 – 15:08) in Australia, 14.1 minutes (IQR: 10.5 – 20.2) in Austria, 13.4 minutes (IQR: 9.8 – 19.0) in Germany, 16.4 (IQR: 12.8 – 27.6) in Hungary, 17.3 (IQR: 12.0 – 22.5) in Italy, 11.2 minutes (IQR: 8.0 – 15.7) in Slovenia and 11.9 minutes (IQR: 9.0 – 18.3) in Switzerland. The completion rate of the survey ranged from 63.3% in Slovenia to 91.7% in Australia (Italy: 66.1%, Hungary: 67.3%, Austria: 79.7%, Germany: 85.2%, Switzerland: 87.8%).

### Statistics

Baseline characteristics are presented as mean ±SD or median (min-max), as appropriate. Categorical variables are provided as absolute numbers and in percent. In the main analysis, environmental variables (country of survey: Germany vs. Austria, size of town of practice (< 5,000 vs. 5,000 - <20,000 vs. 20,000 - <100,000 vs. ≥100,000), type of practice: single-handed vs. not single handed;) that may have influenced responses were analyzed using General Linear Models. The main effects and all two-way interactions were therefore analyzed. Bonferroni correction was used to take account of multiple testing.

Estimated means and 95% confidence intervals were used to present the results. For a better understanding of results, responses to items were also presented. In this presentation, the response categories “yes” and “probably yes” and the response categories “probably no” and “no” were combined. No statistical correction was carried out to adjust for non-representative samples.

### Ethics

The study protocol was approved by the local ethics committee of Goethe University Frankfurt, Germany (ethics committee number 20-619), Bond University, Australia and Slovenia. According to national laws in Austria, Italy, Hungary and Switzerland no approval of the local ethics committee was necessary.

**Appendix Table 3.**
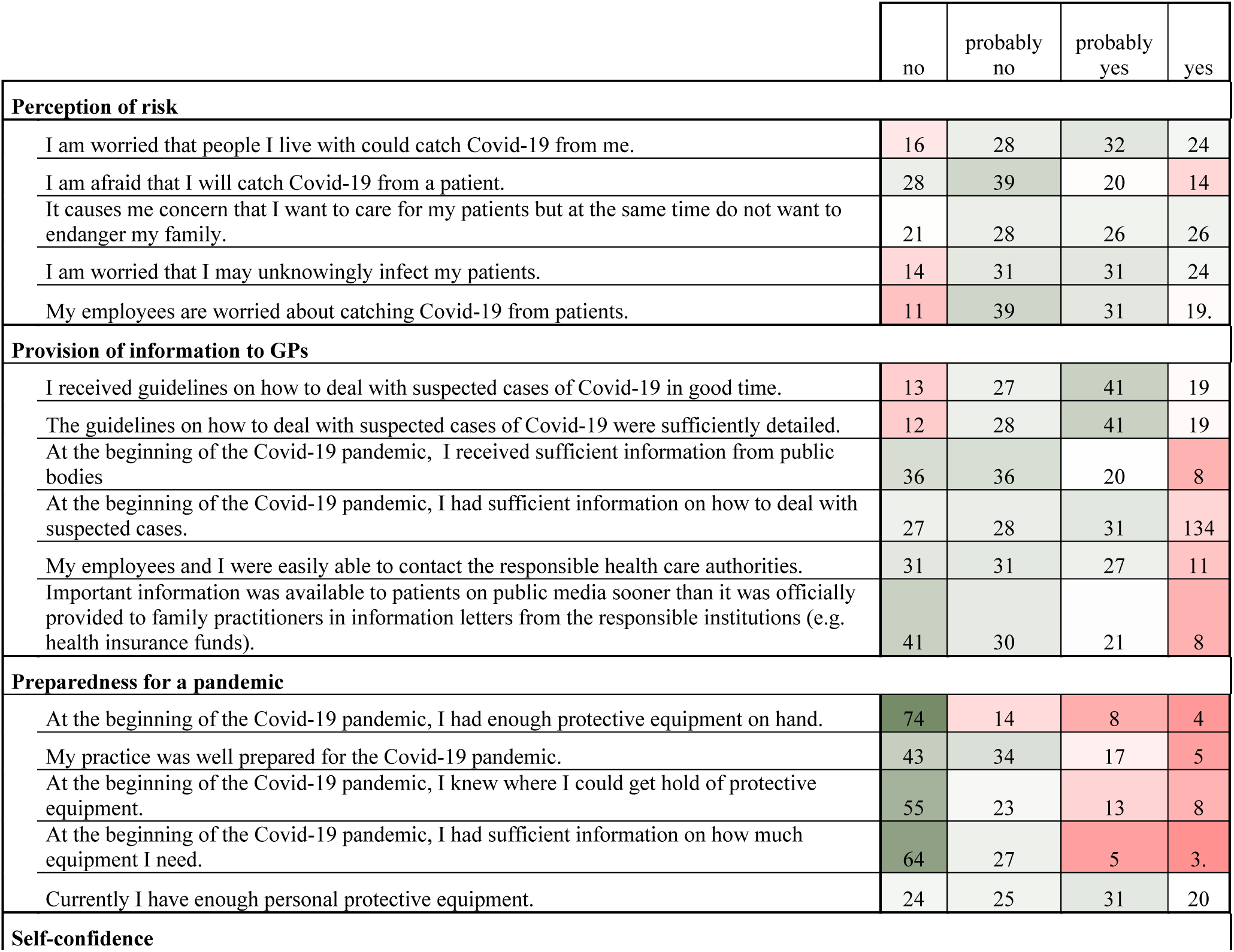
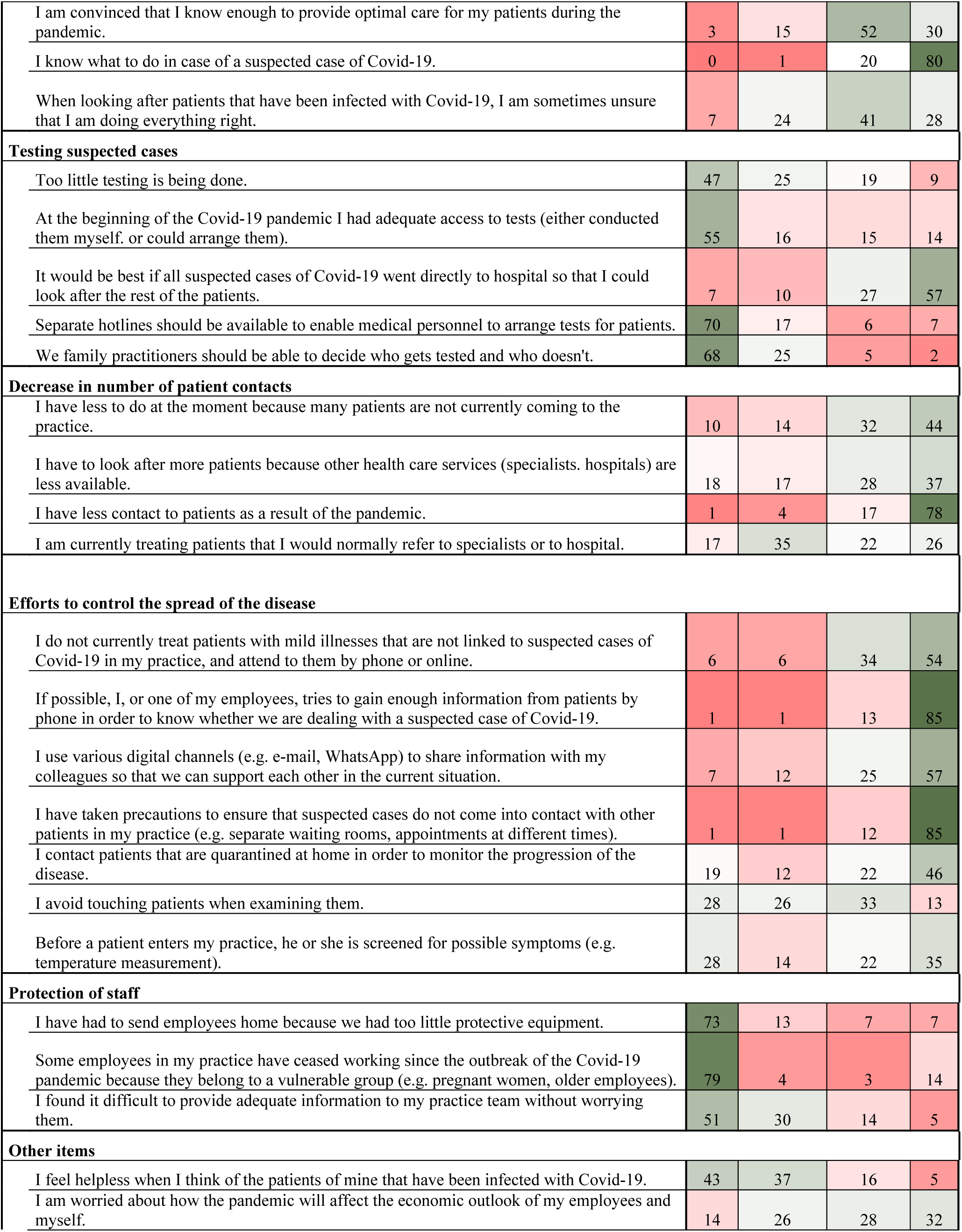
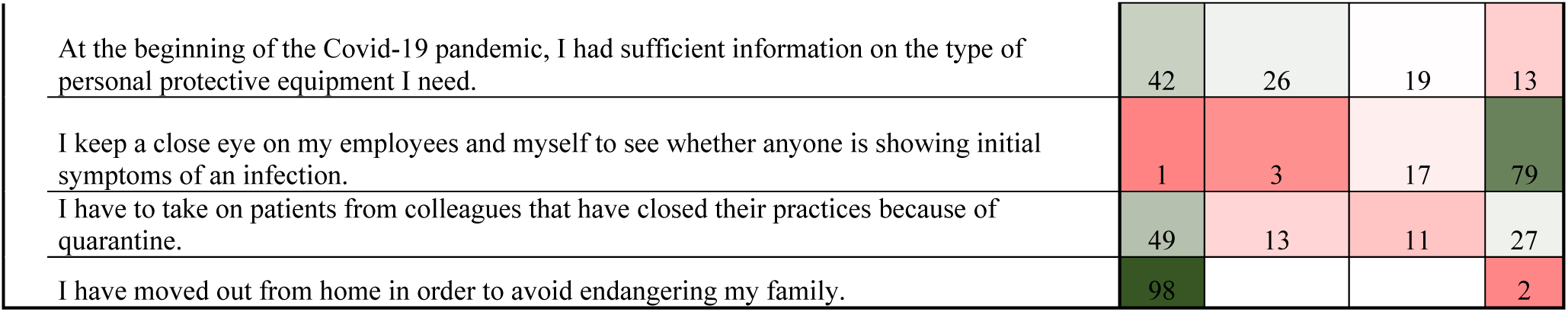
Response distribution (%) for all items

|  | no | probably no | probably yes | yes |
| --- | --- | --- | --- | --- |
| **Perception of risk** |  |  |  |  |
| I am worried that people I live with could catch Covid-19 from me. | 16 | 28 | 32 | 24 |
| I am afraid that I will catch Covid-19 from a patient. | 28 | 39 | 20 | 14 |
| It causes me concern that I want to care for my patients but at the same time do not want to endanger my family. | 21 | 28 | 26 | 26 |
| I am worried that I may unknowingly infect my patients. | 14 | 31 | 31 | 24 |
| My employees are worried about catching Covid-19 from patients. | 11 | 39 | 31 | 19. |
| **Provision of information to GPs** |  |  |  |  |
| I received guidelines on how to deal with suspected cases of Covid-19 in good time. | 13 | 27 | 41 | 19 |
| The guidelines on how to deal with suspected cases of Covid-19 were sufficiently detailed. | 12 | 28 | 41 | 19 |
| At the beginning of the Covid-19 pandemic, I received sufficient information from public bodies | 36 | 36 | 20 | 8 |
| At the beginning of the Covid-19 pandemic, I had sufficient information on how to deal with suspected cases. | 27 | 28 | 31 | 134 |
| My employees and I were easily able to contact the responsible health care authorities. | 31 | 31 | 27 | 11 |
| Important information was available to patients on public media sooner than it was officially provided to family practitioners in information letters from the responsible institutions (e.g. health insurance funds). | 41 | 30 | 21 | 8 |
| **Preparedness for a pandemic** |  |  |  |  |
| At the beginning of the Covid-19 pandemic, I had enough protective equipment on hand. | 74 | 14 | 8 | 4 |
| My practice was well prepared for the Covid-19 pandemic. | 43 | 34 | 17 | 5 |
| At the beginning of the Covid-19 pandemic, I knew where I could get hold of protective equipment. | 55 | 23 | 13 | 8 |
| At the beginning of the Covid-19 pandemic, I had sufficient information on how much equipment I need. | 64 | 27 | 5 | 3. |
| Currently I have enough personal protective equipment. | 24 | 25 | 31 | 20 |
| **Self-confidence** |  |  |  |  |
| I am convinced that I know enough to provide optimal care for my patients during the pandemic. | 3 | 15 | 52 | 30 |
| I know what to do in case of a suspected case of Covid-19. | 0 | 1 | 20 | 80 |
| When looking after patients that have been infected with Covid-19, I am sometimes unsure that I am doing everything right. | 7 | 24 | 41 | 28 |
| **Testing suspected cases** |  |  |  |  |
| Too little testing is being done. | 47 | 25 | 19 | 9 |
| At the beginning of the Covid-19 pandemic I had adequate access to tests (either conducted them myself, or could arrange them). | 55 | 16 | 15 | 14 |
| It would be best if all suspected cases of Covid-19 went directly to hospital so that I could look after the rest of the patients. | 7 | 10 | 27 | 57 |
| Separate hotlines should be available to enable medical personnel to arrange tests for patients. | 70 | 17 | 6 | 7 |
| We family practitioners should be able to decide who gets tested and who doesn't. | 68 | 25 | 5 | 2 |
| **Decrease in number of patient contacts** |  |  |  |  |
| I have less to do at the moment because many patients are not currently coming to the practice. | 10 | 14 | 32 | 44 |
| I have to look after more patients because other health care services (specialists, hospitals) are less available. | 18 | 17 | 28 | 37 |
| I have less contact to patients as a result of the pandemic. | 1 | 4 | 17 | 78 |
| I am currently treating patients that I would normally refer to specialists or to hospital. | 17 | 35 | 22 | 26 |
| **Efforts to control the spread of the disease** |  |  |  |  |
| I do not currently treat patients with mild illnesses that are not linked to suspected cases of Covid-19 in my practice, and attend to them by phone or online. | 6 | 6 | 34 | 54 |
| If possible, I, or one of my employees, tries to gain enough information from patients by phone in order to know whether we are dealing with a suspected case of Covid-19. | 1 | 1 | 13 | 85 |
| I use various digital channels (e.g. e-mail, WhatsApp) to share information with my colleagues so that we can support each other in the current situation. | 7 | 12 | 25 | 57 |
| I have taken precautions to ensure that suspected cases do not come into contact with other patients in my practice (e.g. separate waiting rooms, appointments at different times). | 1 | 1 | 12 | 85 |
| I contact patients that are quarantined at home in order to monitor the progression of the disease. | 19 | 12 | 22 | 46 |
| I avoid touching patients when examining them. | 28 | 26 | 33 | 13 |
| Before a patient enters my practice, he or she is screened for possible symptoms (e.g. temperature measurement). | 28 | 14 | 22 | 35 |
| **Protection of staff** |  |  |  |  |
| I have had to send employees home because we had too little protective equipment. | 73 | 13 | 7 | 7 |
| Some employees in my practice have ceased working since the outbreak of the Covid-19 pandemic because they belong to a vulnerable group (e.g. pregnant women, older employees). | 79 | 4 | 3 | 14 |
| I found it difficult to provide adequate information to my practice team without worrying them. | 51 | 30 | 14 | 5 |
| **Other items** |  |  |  |  |
| I feel helpless when I think of the patients of mine that have been infected with Covid-19. | 43 | 37 | 16 | 5 |
| I am worried about how the pandemic will affect the economic outlook of my employees and myself. | 14 | 26 | 28 | 32 |
| At the beginning of the Covid-19 pandemic, I had sufficient information on the type of personal protective equipment I need. | 42 | 26 | 19 | 13 |
| I keep a close eye on my employees and myself to see whether anyone is showing initial symptoms of an infection. | 1 | 3 | 17 | 79 |
| I have to take on patients from colleagues that have closed their practices because of quarantine. | 49 | 13 | 11 | 27 |
| I have moved out from home in order to avoid endangering my family. | 98 |  |  | 2 |

**Appendix Table 4.** Difference in the responses of Austrian and German GPs. Percentages were calculated as %German GPs minus %Austrian GPs. Responses which were more often chosen by German GPs are marked green and responses which were more often chosen by Austrian GPs are marked red.

|  | no | probably<br>no | probably<br>yes | yes |
| --- | --- | --- | --- | --- |
| **Perception of risk** |  |  |  |  |
| I am worried that people I live with could catch Covid-19 from me. | 0 | -8 | -2 | 10 |
| I am afraid that I will catch Covid-19 from a patient. | 0 | -6 | 1 | 5 |
| It causes me concern that I want to care for my patients but at the same time do not want to endanger my family. | 2 | -5 | -2 | 6 |
| I am worried that I may unknowingly infect my patients. | -2 | -5 | 2 | 6 |
| My employees are worried about catching Covid-19 from patients. | -6 | -8 | 8 | 6 |
| **Provision of information to GPs** |  |  |  |  |
| I received guidelines on how to deal with suspected cases of Covid-19 in good time. | -3 | -3 | 1 | 6 |
| The guidelines on how to deal with suspected cases of Covid-19 were sufficiently detailed. | -4 | -2 | 3 | 4 |
| At the beginning of the Covid-19 pandemic I received sufficient information from public bodies | 0 | 2 | -2 | 0 |
| At the beginning of the Covid-19 pandemic I had sufficient information on how to deal with suspected cases. | -1 | 1 | -1 | 0 |
| My employees and I were easily able to contact the responsible health care authorities. | 8 | 0 | -6 | -2 |
| Important information was available to patients on public media sooner than it was officially provided to family practitioners in information letters from the responsible institutions (e.g. health insurance funds). | 2 | 7 | 0 | -8 |
| **Preparedness for a pandemic** |  |  |  |  |
| At the beginning of the Covid-19 pandemic, I had enough protective equipment on hand. | 1 | 1 | 3 | -5 |
| My practice was well prepared for the Covid-19 pandemic. | -5 | 1 | 3 | 0 |
| At the beginning of the Covid-19 pandemic, I knew where I could get hold of protective equipment. | -8 | 4 | 3 | 1 |
| At the beginning of the Covid-19 pandemic, I had sufficient information on how much equipment I need. | -1 | 0 | 0 | 0 |
| Currently I have enough personal protective equipment. | -4 | -2 | 1 | 5 |
| **Self-confidence** |  |  |  |  |
| I am convinced that I know enough to provide optimal care for my patients during the pandemic. | 1 | 5 | -1 | -5 |
| I know what to do in case of a suspected case of Covid-19. | 0 | -1 | 3 | -2 |
| When looking after patients that have been infected with Covid-19, I am sometimes unsure that I am doing everything right. | -2 | -1 | 2 | 1 |
| **Testing suspected cases** |  |  |  |  |
| Too little testing is being done. | -22 | 0 | 14 | 8 |
| At the beginning of the Covid-19 pandemic, I had adequate access to tests (either conducted them myself, or could arrange them). | 18 | 14 | 6 | -39 |
| It would be best if all suspected cases of Covid-19 went directly to hospital so that I could look after the rest of the patients. | 14 | 0 | -8 | -5 |
| Separate hotlines should be available to enable medical personnel to arrange tests for patients. | -20 | 6 | 5 | 9 |
| We family practitioners should be able to decide who gets tested and who doesn't. | -13 | 8 | 3 | 2 |
| **Decrease in number of patient contacts** |  |  |  |  |
| I have less to do at the moment because many patients are not currently coming to the practice. | -4 | 6 | 2 | -3 |
| I have to look after more patients because other health care services (specialists, hospitals) are less available. | 14 | 4 | -7 | -12 |
| I have less contact to patients as a result of the pandemic. | 0 | 2 | 13 | -15 |
| I am currently treating patients that I would normally refer to specialists or to hospital. | -9 | -6 | 9 | 5 |
| **Efforts to control the spread of the disease** |  |  |  |  |
| I do not currently treat patients with mild illnesses that are not linked to suspected cases of Covid-19 in my practice, and attend to them by phone or online. | 4 | 3 | 7 | -14 |
| If possible, I, or one of my employees, tries to gain enough information from patients by phone in order to know whether we are dealing with a suspected case of Covid-19. | 0 | -1 | 5 | -4 |
| I use various digital channels (e.g. e-mail, WhatsApp) to share information with my colleagues so that we can support each other in the current situation. | 1 | 5 | 4 | -10 |
| I have taken precautions to ensure that suspected cases do not come into contact with other patients in my practice (e.g. separate waiting rooms, appointments at different times). | 0 | 1 | 5 | -6 |
| I contact patients that are quarantined at home in order to monitor the progression of the disease. | -13 | -1 | 11 | 2 |
| I avoid touching patients when examining them. | 5 | 3 | -3 | -4 |
| Before a patient enters my practice, he or she is screened for possible symptoms (e.g. temperature measurement). | 4 | 2 | 2 | -8 |
| **Protection of staff** |  |  |  |  |
| I have had to send employees home because we had too little protective equipment. | 8 | 0 | -3 | -4 |
| Some employees in my practice have ceased working since the outbreak of the Covid-19 pandemic because they belong to a vulnerable group (e.g. pregnant women, older employees). | -2 | 3 | 1 | -2 |
| I found it difficult to provide adequate information to my practice team without worrying them. | -7 | 1 | 5 | 1 |
| **Other items** |  |  |  |  |
| I feel helpless when I think of the patients of mine that have been infected with Covid-19. | 1 | 0 | 1 | -1 |
| I am worried about how the pandemic will affect the economic outlook of my employees and myself. | -5 | -5 | 4 | 6 |
| At the beginning of the Covid-19 pandemic, I had sufficient information on the type of personal protective equipment I need. | -6 | -1 | 4 | 2 |
| I keep a close eye on my employees and myself to see whether anyone is showing initial symptoms of an infection. | 1 | 4 | 6 | -10 |
| I have to take on patients from colleagues that have closed their practices because of quarantine. | 8 | 1 | -1 | -8 |
| I have moved out from home in order to avoid endangering my family. | 1 | -1 | 0 | 0 |

**Appendix Figure 3.**
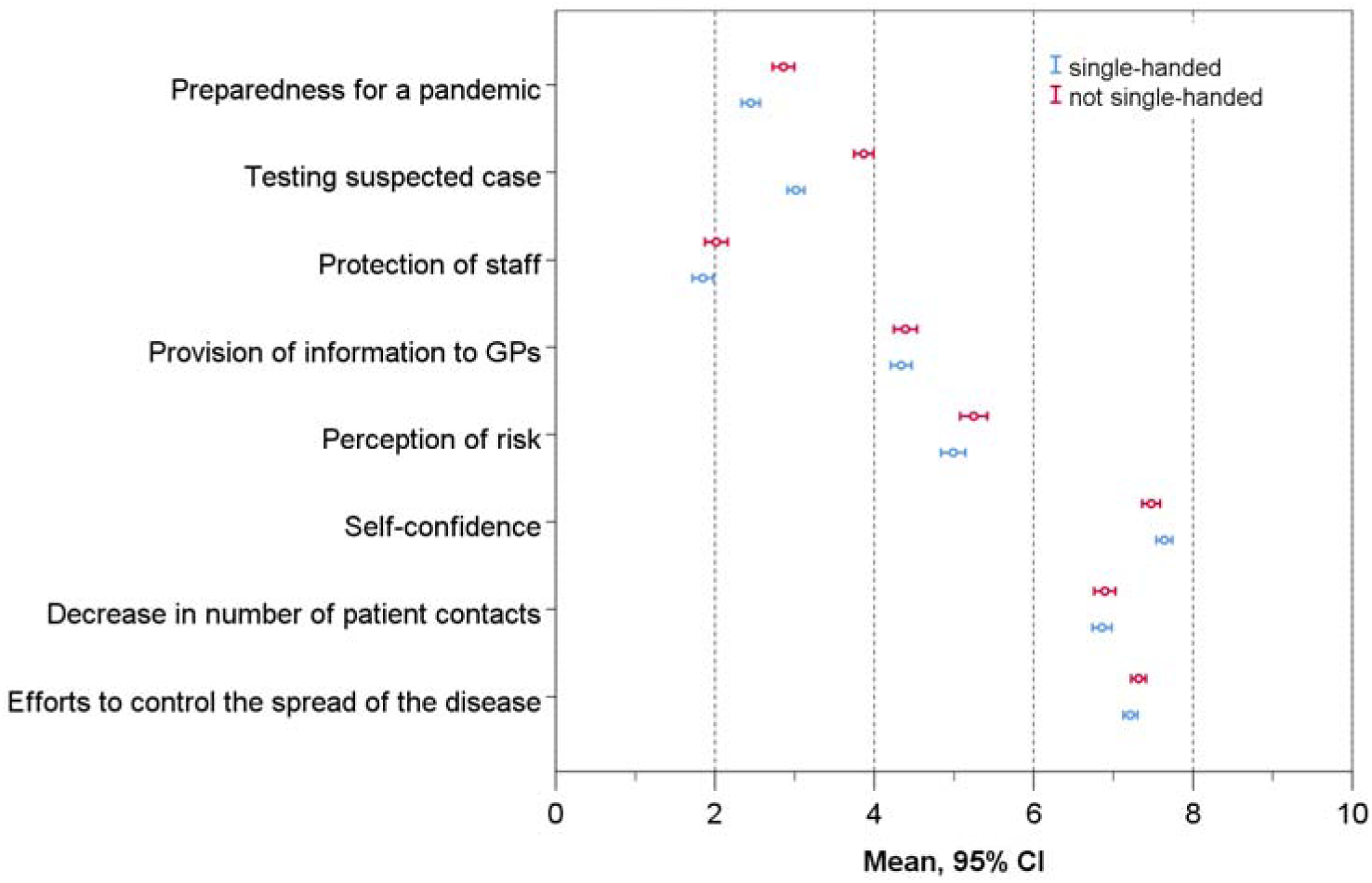
Differences between GPs in single-handed and not single-handed practices in their evaluation of the pandemic.

Checklist for Reporting Results of Internet E-Surveys (CHERRIES)

(1) COVI-Prim Baseline Questionarie - German

Checklist for Reporting Results of Internet E-Surveys (CHERRIES)
| *Item Category* | *Checklist Item* | *Page* |
| --- | --- | --- |
| **Design** | Describe survey design | 6 |
| **IRB (Institutional Review Board) approval and informed consent process** | IRB approval | 6 |
|  | Informed consent | A-2, A-7 |
|  | Data protection | A-7 |
| **Development and pre-testing** | Development and testing | 5, A-2 |
| **Recruitment process and description of the sample having access to the questionnaire** | Open survey versus closed survey | A-5 |
|  | Contact mode | A-6 |
|  | Advertising the survey | A-6 |
| **Survey administration** | Web/E-mail | 6, A-5 |
|  | Context | A-5 |
|  | Mandatory/voluntary | A-5 |
|  | Incentives | A-5 |
|  | Time/Date | A-5 |
|  | Randomization of items or questionnaires | A-6 |
|  | Adaptive questioning | A-6 |
|  | Number of Items | 5, A-2, A-3, A-4 |
|  | Number of screens (pages) | A-3, A-4 |
|  | Completeness check | A-6 |
|  | Review step | A-6 |
| **Response rates** | Unique site visitor | NA |
|  | View rate (Ratio of unique survey visitors/unique site visitors) | NA |
|  | Participation rate (Ratio of unique visitors who agreed to participate/unique first survey page visitors) | NA |
|  | Completion rate (Ratio of users who finished the survey/users who agreed to participate) | 2,7, A-7 |
| **Preventing multiple entries from the same individual** | Cookies used | A-6 |
|  | IP check | A-6 |
|  | Log file analysis | A-6 |
|  | Registration | NA |
| **Analysis** | Handling of incomplete questionnaires | A-4 |
|  | Questionnaires submitted with an atypical timestamp | A-6 |
|  | Statistical correction | A-7 |

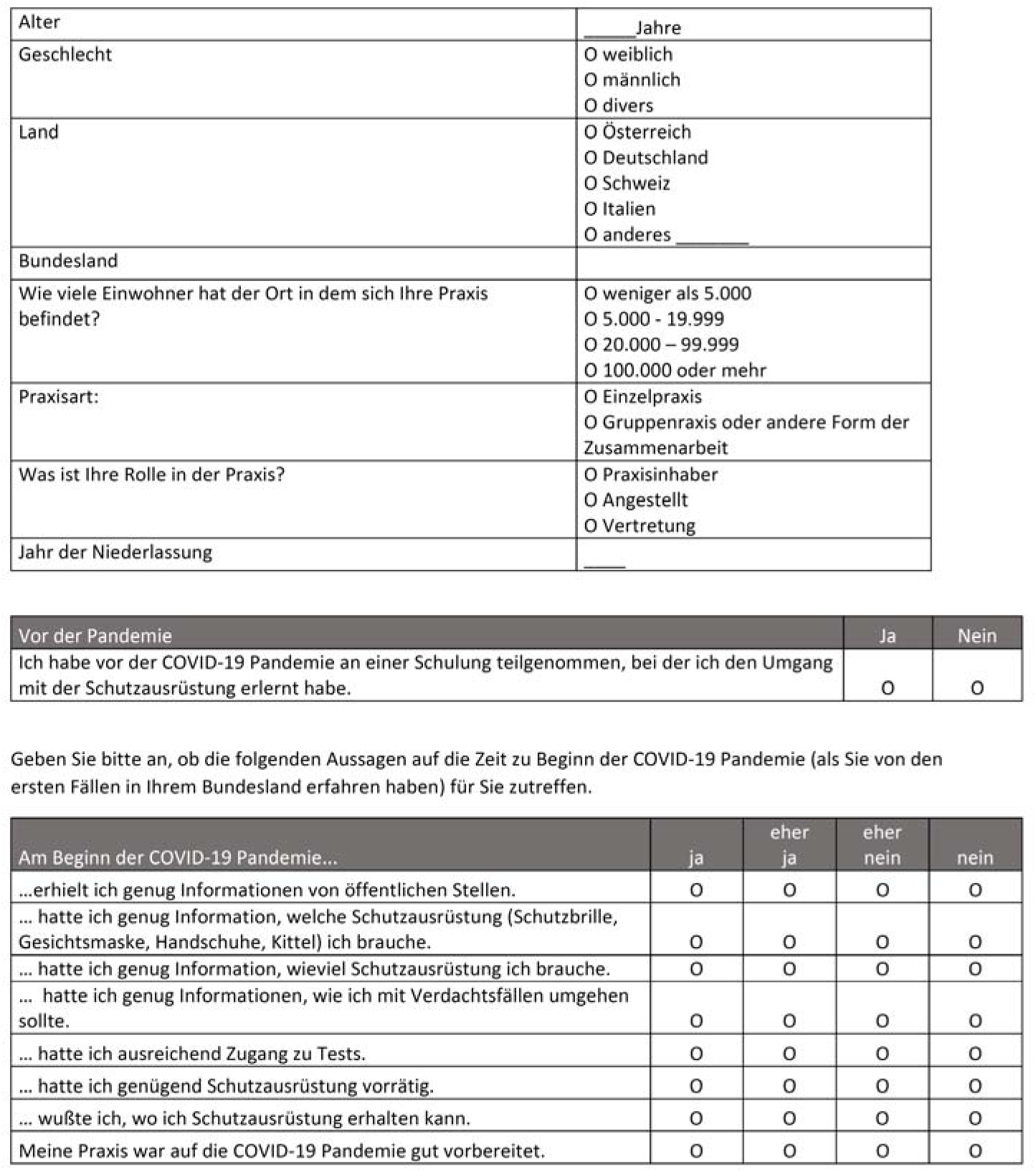

|  |  |
| --- | --- |
| Alter | _____ Jahre |
| Geschlecht | ☐ weiblich<br>☐ männlich<br>☐ divers |
| Land | ☐ Österreich<br>☐ Deutschland<br>☐ Schweiz<br>☐ Italien<br>☐ anderes _____ |
| Bundesland | _____ |
| Wie viele Einwohner hat der Ort in dem sich Ihre Praxis befindet? | ☐ weniger als 5.000<br>☐ 5.000 - 19.999<br>☐ 20.000 – 99.999<br>☐ 100.000 oder mehr |
| Praxisart: | ☐ Einzelpraxis<br>☐ Gruppenpraxis oder andere Form der Zusammenarbeit |
| Was ist Ihre Rolle in der Praxis? | ☐ Praxisinhaber<br>☐ Angestellt<br>☐ Vertretung |
| Jahr der Niederlassung | _____ |

| Vor der Pandemie | Ja | Nein |
| --- | --- | --- |
| Ich habe vor der COVID-19 Pandemie an einer Schulung teilgenommen, bei der ich den Umgang mit der Schutzausrüstung erlernt habe. | ☐ | ☐ |

| Am Beginn der COVID-19 Pandemie... | ja | eher ja | eher nein | nein |
| --- | --- | --- | --- | --- |
| ...erhielt ich genug Informationen von öffentlichen Stellen. | ☐ | ☐ | ☐ | ☐ |
| ... hatte ich genug Information, welche Schutzausrüstung (Schutzbrille, Gesichtsmaske, Handschuhe, Kittel) ich brauche. | ☐ | ☐ | ☐ | ☐ |
| ... hatte ich genug Information, wieviel Schutzausrüstung ich brauche. | ☐ | ☐ | ☐ | ☐ |
| ... hatte ich genug Informationen, wie ich mit Verdachtsfällen umgehen sollte. | ☐ | ☐ | ☐ | ☐ |
| ... hatte ich ausreichend Zugang zu Tests. | ☐ | ☐ | ☐ | ☐ |
| ... hatte ich genügend Schutzausrüstung vorrätig. | ☐ | ☐ | ☐ | ☐ |
| ... wußte ich, wo ich Schutzausrüstung erhalten kann. | ☐ | ☐ | ☐ | ☐ |
| Meine Praxis war auf die COVID-19 Pandemie gut vorbereitet. | ☐ | ☐ | ☐ | ☐ |

| Informationsfluss während der Pandemie | ja | eher ja | eher nein | nein |
| --- | --- | --- | --- | --- |
| Die Richtlinien zum Umgang mit COVID-19 Verdachtsfällen erhielt ich rechtzeitig. | ☐ | ☐ | ☐ | ☐ |
| Die Richtlinien zum Umgang mit COVID-19 Verdachtsfällen waren ausreichend detailliert. | ☐ | ☐ | ☐ | ☐ |
| Verantwortliche Stellen im Gesundheitswesen waren für mich immer gut erreichbar. | ☐ | ☐ | ☐ | ☐ |

| Umgang mit der Pandemie | ja | eher ja | eher nein | nein |
| --- | --- | --- | --- | --- |
| Ich tausche mich über unterschiedliche digitale Kanäle (z.B. E-Mail, WhatsApp) mit meinen KollegInnen aus, um uns in der derzeitigen Situation zu unterstützen. | ☐ | ☐ | ☐ | ☐ |
| * Derzeit habe ich genügend Schutzausrüstung. | ☐ | ☐ | ☐ | ☐ |
| Ich musste MitarbeiterInnen nach Hause schicken, da wir zu wenig Schutzausrüstung hatten. | ☐ | ☐ | ☐ | ☐ |
| Ich beobachte meine MitarbeiterInnen und mich genau, ob jemand erste Symptome einer Infektion zeigt. | ☐ | ☐ | ☐ | ☐ |
| Einzelne MitarbeiterInnen meiner Praxis arbeiten seit der COVID-19 Pandemie nicht mehr, da sie einer sensiblen Gruppe angehören (z.B. Schwangere, ältere MitarbeiterInnen) | ☐ | ☐ | ☐ | ☐ |
| Für mich war es schwierig mein Team ausreichend zu informieren, ohne die MitarbeiterInnen zu sehr zu ängstigen. | ☐ | ☐ | ☐ | ☐ |
| Wenn möglich versuche ich telefonisch genügend Informationen von der/dem PatientIn zu erhalten, um zu wissen, ob es sich um einen COVID-19 Verdachtsfall handelt. | ☐ | ☐ | ☐ | ☐ |
| Bevor ein/e PatientIn meine Praxis betritt, wird er/sie auf mögliche Symptome gescreent. (z.B. Fiebermessung) | ☐ | ☐ | ☐ | ☐ |
| Ich habe Vorkehrungen getroffen, damit sich Verdachtsfälle und andere PatientInnen in meiner Praxis nicht begegnen (z.B. getrennte Warteräume, Termine zu unterschiedlichen Uhrzeiten) | ☐ | ☐ | ☐ | ☐ |

| Umgang mit der Pandemie | ja | eher ja | eher nein | nein |
| --- | --- | --- | --- | --- |
| *PatientInnen mit leichteren Erkrankungen behandle ich derzeit nicht in meiner Praxis, sondern betreue sie am Telefon oder online. | 0 | 0 | 0 | 0 |
| Ich vermeide es, PatientInnen bei der Untersuchung zu berühren. | 0 | 0 | 0 | 0 |
| *Durch die Pandemie habe ich viel weniger PatientInnenkontakte. | 0 | 0 | 0 | 0 |
| *Ich kontaktiere PatientInnen, die in Heimquarantäne sind, um den Verlauf der Erkrankung zu überwachen. | 0 | 0 | 0 | 0 |
| *Ich muss PatientInnen von KollegInnen übernehmen, die ihre Praxis wegen einer Quarantäne geschlossen haben. | 0 | 0 | 0 | 0 |
| *Ich muss mich um mehr PatientInnen kümmern, da andere Versorgungsstrukturen (Fachärzte, Krankenhäuser) weniger verfügbar sind. | 0 | 0 | 0 | 0 |
| * Da viele PatientInnen derzeit nicht in die Praxis kommen, habe ich derzeit weniger zu tun. | 0 | 0 | 0 | 0 |
| *Ich versorge derzeit mehr PatientInnen selbst, die ich normalerweise überweisen würde. | 0 | 0 | 0 | 0 |
| Für medizinisches Personal sollte es eigene Hotlines geben, um Tests für PatientInnen veranlassen zu können. | 0 | 0 | 0 | 0 |
| Es wird zu wenig getestet. | 0 | 0 | 0 | 0 |
| Wir als HausärztInnen sollten entscheiden können, wer getestet wird und wer nicht getestet wird. | 0 | 0 | 0 | 0 |
| Ich bin überzeugt, dass ich genug weiß, um meine PatientInnen optimal während dieser Pandemie zu betreuen. | 0 | 0 | 0 | 0 |
| Ich weiß, was ich bei einem COVID-19 Verdachtsfall tun muss. | 0 | 0 | 0 | 0 |
| Wichtige Informationen waren für die Öffentlichkeit über Medien früher verfügbar, als für uns HausärztInnen über offizielle Informationsschreiben zuständiger Einrichtungen. | 0 | 0 | 0 | 0 |
| Am besten wäre es, wenn alle COVID-19 Verdachtsfälle direkt ins Krankenhaus gehen, damit ich mich um die restlichen PatientInnen kümmern kann. | 0 | 0 | 0 | 0 |

| Personal worries | ja | eher ja | eher nein | nein |
| --- | --- | --- | --- | --- |
| Es belastet mich, dass ich einerseits meine PatientInnen betreuen will, aber auch meine Familie nicht gefährden möchte. | 0 | 0 | 0 | 0 |
| *Ich habe Angst, dass ich mich bei einem/r PatientIn mit COVID-19 infiziere. | 0 | 0 | 0 | 0 |

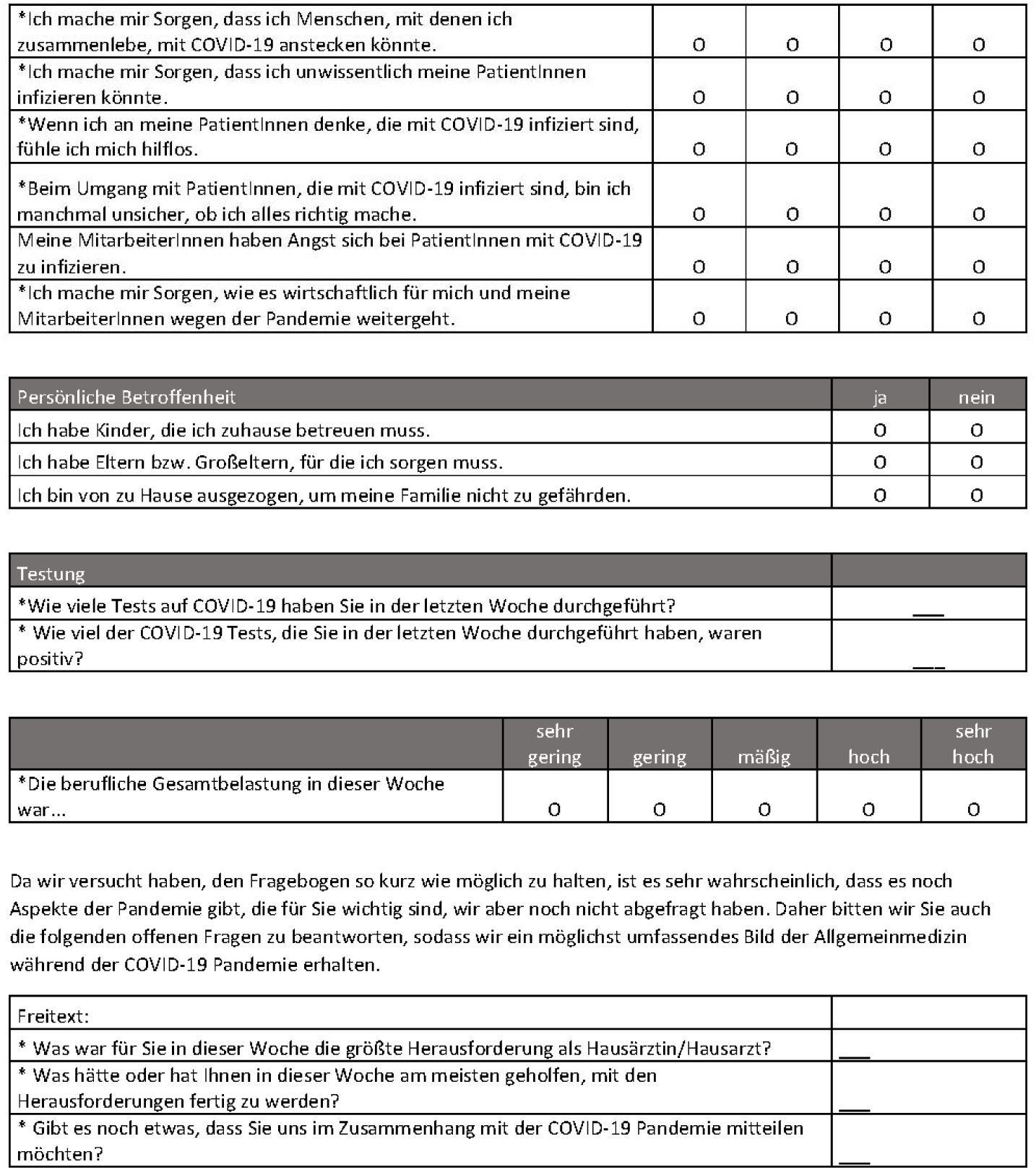

|  |  |  |  |  |
| --- | --- | --- | --- | --- |
| *Ich mache mir Sorgen, dass ich Menschen, mit denen ich zusammenlebe, mit COVID-19 anstecken könnte. | 0 | 0 | 0 | 0 |
| *Ich mache mir Sorgen, dass ich unwissentlich meine PatientInnen infizieren könnte. | 0 | 0 | 0 | 0 |
| *Wenn ich an meine PatientInnen denke, die mit COVID-19 infiziert sind, fühle ich mich hilflos. | 0 | 0 | 0 | 0 |
| *Beim Umgang mit PatientInnen, die mit COVID-19 infiziert sind, bin ich manchmal unsicher, ob ich alles richtig mache. | 0 | 0 | 0 | 0 |
| Meine MitarbeiterInnen haben Angst sich bei PatientInnen mit COVID-19 zu infizieren. | 0 | 0 | 0 | 0 |
| *Ich mache mir Sorgen, wie es wirtschaftlich für mich und meine MitarbeiterInnen wegen der Pandemie weitergeht. | 0 | 0 | 0 | 0 |

| Persönliche Betroffenheit | ja | nein |
| --- | --- | --- |
| Ich habe Kinder, die ich zuhause betreuen muss. | 0 | 0 |
| Ich habe Eltern bzw. Großeltern, für die ich sorgen muss. | 0 | 0 |
| Ich bin von zu Hause ausgezogen, um meine Familie nicht zu gefährden. | 0 | 0 |

| Testung |  |
| --- | --- |
| *Wie viele Tests auf COVID-19 haben Sie in der letzten Woche durchgeführt? | ___ |
| * Wie viel der COVID-19 Tests, die Sie in der letzten Woche durchgeführt haben, waren positiv? | ___ |

|  | sehr gering | gering | mäßig | hoch | sehr hoch |
| --- | --- | --- | --- | --- | --- |
| *Die berufliche Gesamtbelastung in dieser Woche war... | 0 | 0 | 0 | 0 | 0 |

|  |  |
| --- | --- |
| Freitext: |  |
| * Was war für Sie in dieser Woche die größte Herausforderung als Hausärztin/Hausarzt? | ___ |
| * Was hätte oder hat Ihnen in dieser Woche am meisten geholfen, mit den Herausforderungen fertig zu werden? | ___ |
| * Gibt es noch etwas, dass Sie uns im Zusammenhang mit der COVID-19 Pandemie mitteilen möchten? | ___ |

|  |  |
| --- | --- |
| *Wie viel Stunden haben Sie in der letzten Woche gearbeitet? | ___ |
| *Gemessen an Ihrer Gesamtarbeitszeit... |  |
| *...wie groß war der Anteil an Telefonkonsultationen? | ___ % |
| *...wie groß war der Anteil an Konsultationen in der Praxis? | ___ % |
| *...wie groß war der Anteil an Koordinations und Organisationstätigkeit? | ___ % |
| *...wie groß war der Anteil an anderen, nicht genannten Tätigkeiten? | ___ % |
| *Welche anderen Tätigkeiten sind dies? | ___ |
| *Gemessen an Ihrer Gesamtarbeitszeit... |  |
| *Wie viel Ihrer gesamten Arbeitszeit hat derzeit direkt oder indirekt mit COVID-19 zu tun? | ___ % |
| Wie viel Ihrer gesamten Arbeitszeit entfällt derzeit auf übliche Versorgung wie Vorsorge oder Betreuung chronisch kranker Personen? | ___ % |
| *Wie viel Ihrer gesamten Arbeitszeit entfällt auf andere, nicht genannte Tätigkeiten? | ___ % |
| *Welche anderen Tätigkeiten sind dies? | ___ |

\* Diese Items sind Teil der wöchentlichen Befragung.

(1) COVI-Prim Baseline Questionarie - English

|  |  |
| --- | --- |
| Age (years) | _____ Years |
| Sex | ☐ female<br>☐ male<br>☐ diverse |
| Country | ☐ Australia<br>☐ other _____ |
| State |  |
| How many inhabitants are there in the municipality where your practice is located | ☐ fewer than 5.000<br>☐ 5.000 - 19.999<br>☐ 20.000 - 99.999<br>☐ 100.000 or more |
| Type of practice: | ☐ single-handed<br>☐ not single-handed |
| What is your position in the practice? | ☐ owner<br>☐ employed<br>☐ substitute |
| How long has the practice existed? | _____ Years |

| Prior to the pandemic | Yes | No |
| --- | --- | --- |
| Prior to the pandemic, I took part in a training course in which I learned how to use personal protective equipment (protective eyewear, face mask, glasses, gloves, coats). | ☐ | ☐ |

| At the beginning of the Covid-19 pandemic... | Yes | probably yes | probably no | No |
| --- | --- | --- | --- | --- |
| ... I received sufficient information from public bodies | ☐ | ☐ | ☐ | ☐ |
| ... I had sufficient information on the type of personal protective equipment I need. | ☐ | ☐ | ☐ | ☐ |
| ... I had sufficient information on how much equipment I need. | ☐ | ☐ | ☐ | ☐ |
| ... I had sufficient information on how to deal with suspected cases. | ☐ | ☐ | ☐ | ☐ |
| ... I had adequate access to tests (either conducted them myself, or could arrange them). | ☐ | ☐ | ☐ | ☐ |
| ... I had enough protective equipment on hand. | ☐ | ☐ | ☐ | ☐ |
| ... I knew where I could get hold of protective equipment. | ☐ | ☐ | ☐ | ☐ |
| My practice was well prepared for the Covid-19 pandemic. | ☐ | ☐ | ☐ | ☐ |

| Flow of information during the pandemic | Yes | probably yes | probably no | No |
| --- | --- | --- | --- | --- |
| I received guidelines on how to deal with suspected cases of Covid-19 in good time. | ☐ | ☐ | ☐ | ☐ |
| The guidelines on how to deal with suspected cases of Covid-19 were sufficiently detailed. | ☐ | ☐ | ☐ | ☐ |
| My employees and I were easily able to contact the responsible health care authorities. | ☐ | ☐ | ☐ | ☐ |

| Dealing with the pandemic | Yes | probably yes | probably no | No |
| --- | --- | --- | --- | --- |
| I use various digital channels (e.g. e-mail, WhatsApp) to share information with my colleagues so that we can support each other in the current situation. | ☐ | ☐ | ☐ | ☐ |
| *Currently I have enough personal protective equipment. | ☐ | ☐ | ☐ | ☐ |
| I have had to send employees home because we had too little protective equipment. | ☐ | ☐ | ☐ | ☐ |
| I keep a close eye on my employees and myself to see whether anyone is showing initial symptoms of an infection. | ☐ | ☐ | ☐ | ☐ |
| Some employees in my practice have ceased working since the outbreak of the Covid-19 pandemic because they belong to a vulnerable group (e.g. pregnant women, older employees). | ☐ | ☐ | ☐ | ☐ |
| I found it difficult to provide adequate information to my practice team without worrying them. | ☐ | ☐ | ☐ | ☐ |
| If possible, I, or one of my employees, tries to gain enough information from patients by phone in order to know whether we are dealing with a suspected case of Covid-19. | ☐ | ☐ | ☐ | ☐ |
| Before a patient enters my practice, he or she is screened for possible symptoms (e.g. temperature measurement). | ☐ | ☐ | ☐ | ☐ |
| I have taken precautions to ensure that suspected cases do not come into contact with other patients in my practice (e.g. separate waiting rooms, appointments at different times). | ☐ | ☐ | ☐ | ☐ |
| *I do not currently treat patients with mild illnesses that are not linked to suspected cases of Covid-19 in my practice, and attend to them by phone or online. | ☐ | ☐ | ☐ | ☐ |
| I avoid touching patients when examining them. | ☐ | ☐ | ☐ | ☐ |
| *I have less contact to patients as a result of the pandemic. | ☐ | ☐ | ☐ | ☐ |
| *I contact patients that are quarantined at home in order to monitor the progression of the disease. | ☐ | ☐ | ☐ | ☐ |
| *I have to take on patients from colleagues that have closed their practices because of quarantine. | ☐ | ☐ | ☐ | ☐ |
| *I have to look after more patients because other health care services (specialists, hospitals) are less available. | ☐ | ☐ | ☐ | ☐ |
| *I have less to do at the moment because many patients are not currently coming to the practice. | ☐ | ☐ | ☐ | ☐ |
| *I am currently treating patients that I would normally refer to specialists or to hospital. | ☐ | ☐ | ☐ | ☐ |

| Dealing with the pandemic | Yes | probably yes | probably no | No |
| --- | --- | --- | --- | --- |
| Separate hotlines should be available to enable medical personnel to arrange tests for patients. | ☐ | ☐ | ☐ | ☐ |
| Too little testing is being done. | ☐ | ☐ | ☐ | ☐ |
| We family practitioners should be able to decide who gets tested and who doesn't. | ☐ | ☐ | ☐ | ☐ |
| I am convinced that I know enough to provide optimal care for my patients during the pandemic. | ☐ | ☐ | ☐ | ☐ |
| I know what to do in case of a suspected case of Covid-19. | ☐ | ☐ | ☐ | ☐ |
| Important information was available to patients on public media sooner than it was officially provided to family practitioners in information letters from the responsible institutions (e.g. health insurance funds). | ☐ | ☐ | ☐ | ☐ |
| It would be best if all suspected cases of Covid-19 went directly to hospital so that I could look after the rest of the patients. | ☐ | ☐ | ☐ | ☐ |

| Personal worries | Yes | probably yes | probably no | No |
| --- | --- | --- | --- | --- |
| It causes me concern that I want to care for my patients but at the same time do not want to endanger my family. | ☐ | ☐ | ☐ | ☐ |
| *I am afraid that I will catch Covid-19 from a patient. | ☐ | ☐ | ☐ | ☐ |
| *I am worried that people I live with could catch Covid-19 from me. | ☐ | ☐ | ☐ | ☐ |
| *I am worried that I may unknowingly infect my patients. | ☐ | ☐ | ☐ | ☐ |
| *I feel helpless when I think of the patients of mine that have been infected with Covid-19. | ☐ | ☐ | ☐ | ☐ |
| *When looking after patients that have been infected with Covid-19, I am sometimes unsure that I am doing everything right. | ☐ | ☐ | ☐ | ☐ |
| My employees are worried about catching Covid-19 from patients. | ☐ | ☐ | ☐ | ☐ |
| *I am worried about how the pandemic will affect the economic outlook of my employees and myself. | ☐ | ☐ | ☐ | ☐ |

| Personal concern | Yes | No |
| --- | --- | --- |
| I have children that I have to look after at home. | ☐ | ☐ |
| I have parents and / or grandparents that I have to look after. | ☐ | ☐ |
| I have moved out from home in order to avoid endangering my family. | ☐ | ☐ |

| Personal affect |  |
| --- | --- |
| *How many Covid-19 tests have you performed last week? |  |
| *How many of these Covid-19 tests were positive? |  |

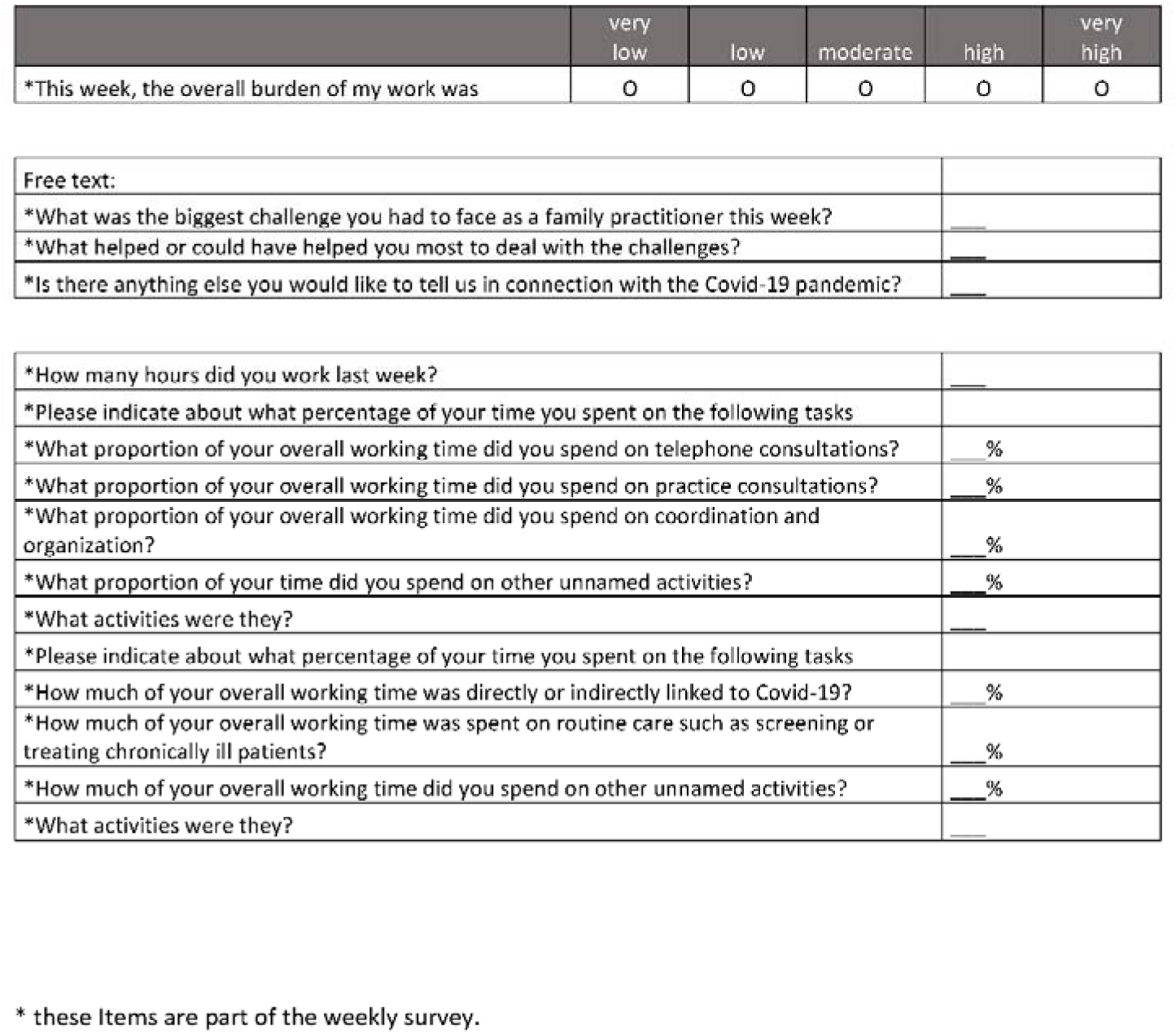

|  | very low | low | moderate | high | very high |
| --- | --- | --- | --- | --- | --- |
| *This week, the overall burden of my work was | 0 | 0 | 0 | 0 | 0 |

|  |  |
| --- | --- |
| Free text: |  |
| *What was the biggest challenge you had to face as a family practitioner this week? | _____ |
| *What helped or could have helped you most to deal with the challenges? | _____ |
| *Is there anything else you would like to tell us in connection with the Covid-19 pandemic? | _____ |
| *How many hours did you work last week? | _____ |
| *Please indicate about what percentage of your time you spent on the following tasks |  |
| *What proportion of your overall working time did you spend on telephone consultations? | _____ % |
| *What proportion of your overall working time did you spend on practice consultations? | _____ % |
| *What proportion of your overall working time did you spend on coordination and organization? | _____ % |
| *What proportion of your time did you spend on other unnamed activities? | _____ % |
| *What activities were they? | _____ |
| *Please indicate about what percentage of your time you spent on the following tasks |  |
| *How much of your overall working time was directly or indirectly linked to Covid-19? | _____ % |
| *How much of your overall working time was spent on routine care such as screening or treating chronically ill patients? | _____ % |
| *How much of your overall working time did you spend on other unnamed activities? | _____ % |
| *What activities were they? | _____ |

\* these items are part of the weekly survey.

